# Subtyping strokes using blood-based biomarkers: A proteomics approach

**DOI:** 10.1101/2023.06.10.23291233

**Authors:** Shubham Misra, Praveen Singh, Shantanu Sengupta, Manoj Kushwaha, Zuhaibur Rahman, Divya Bhalla, Pumanshi Talwar, Manabesh Nath, Rahul Chakraborty, Pradeep Kumar, Amit Kumar, Praveen Aggarwal, Achal K Srivastava, Awadh K Pandit, Dheeraj Mohania, Kameshwar Prasad, Deepti Vibha

## Abstract

**Background and Objectives:** Rapid diagnosis of stroke and its subtypes is critical in early stages. We aimed to discover and validate blood-based protein biomarkers to differentiate ischemic stroke (IS) from intracerebral hemorrhage (ICH) within 24 hours using high-throughput proteomics.

**Methods:** We collected serum samples within 24 hours from acute stroke (IS & ICH) and mimics patients. In the discovery phase, SWATH-MS proteomics identified differentially expressed proteins (fold change: 1.5, p<0.05, and confirmed/tentative selection using Boruta random forest) between IS and ICH which were validated using Multiple Reaction Monitoring (MRM) proteomics in the validation phase. Protein-protein interactions and pathway analysis were conducted using STRING version 11 and Cytoscape 3.9.0. Cut-off points were determined using Youden Index. Prediction models were developed using backward stepwise multivariable logistic regression analysis. Hanley-McNeil test, Integrated discrimination improvement index, and likelihood ratio test determined the improved discrimination ability of biomarkers added to clinical models.

**Results:** Discovery phase included 20 IS and 20 ICH while validation phase included 150 IS, 150 ICH, and 6 stroke mimics. We quantified 365 proteins in the discovery phase. Between IS and ICH, we identified 20 differentially expressed proteins. In the validation phase, combined prediction model including three biomarkers: GFAP (OR 0.04; 95%CI 0.02-0.11), MMP9 (OR 0.09; 95%CI 0.03-0.28), APO-C1 (OR 5.76; 95%CI 2.66-12.47) and clinical variables independently differentiated IS from ICH (accuracy: 92%, sensitivity: 96%, specificity: 69%). Addition of biomarkers to clinical variables improved the discrimination capacity by 26% (p<0.001). Subgroup analysis within 6 hours identified that GFAP and MMP9 differentiated IS from ICH with a sensitivity> 90%.

**Conclusions:** Our study identified that GFAP, MMP, and APO-C1 biomarkers independently differentiated IS from ICH within 24 hours and significantly improved the discrimination ability to predict IS. Temporal profiling of these biomarkers in the acute phase of stroke is urgently warranted.

## Introduction

An early diagnosis of stroke and definitive acute treatment is critical in acute stroke care.^1^ Early detection of stroke and its subtypes is crucial since the efficacy of reperfusion therapies in ischemic stroke (IS) including intravenous thrombolysis^2–4^ and mechanical thrombectomy,^5,6^ is time-dependent. There are several challenges in the diagnosis of IS including poor sensitivity of a computed tomography (CT) scan to diagnose IS in the acute phase, lack of availability of a magnetic resonance imaging (MRI) scan and trained neurologists in resource-limited settings, and expensive cost of imaging equipment’s. Therefore, the search for a novel molecular marker for stroke diagnosis has received growing attention in the last 20 years.^7–9^

As of 2022, we do not have a functional blood-based stroke biomarker (either individual or in combination) which can differentiate IS from intracerebral hemorrhage (ICH) and other similar conditions. We also do not have a point of care test which can strengthen the stroke diagnosis along with neuroimaging. Most of the studies conducted till date have used the candidate-based immunoassays with every protein requiring a separate assay or a multiplex assay approach to determine diagnostic biomarkers for differentiating IS from healthy controls, stroke mimics, and ICH within 24 hours. A high-throughput proteomics approach, on the other hand, can simultaneously quantify many proteins in an exploratory fashion and could potentially be a highly sensitive method to detect diagnostic protein markers in stroke patients at a low cost per sample in a rapid and reproducible manner. Further, recent advancements in label-free quantification of proteins have offered a cost-effective alternative to the labelling methods for discovering protein biomarkers in stroke with subsequent validation in larger cohorts using the targeted proteomics methods. Label-free techniques like sequential windowed acquisition of all theoretical fragment ion mass spectra (SWATH-MS) have been preferred over labelled proteomics techniques for large clinical studies focussed on biomarker discovery.^10^

With the growing need for a blood-based diagnostic biomarker in stroke, it is imperative to utilize the advantages of the latest high-throughput proteomics approaches and focus on relevant clinical questions for identifying protein biomarkers that have adequate sensitivity and/or specificity to differentiate stroke subtypes in clinical settings. Therefore, we aimed to discover blood-based protein biomarkers to diagnose and differentiate IS from ICH within 24 hours using the untargeted SWATH-MS proteomics approach. We also validated the diagnostic performance of the discovered proteins in a separate cohort of IS, ICH, and stroke mimics using the targeted Multiple Reaction Monitoring (MRM) proteomics approach.

## Methods

### Study population and study design

This diagnostic test study was conducted in the Department of Neurology, All India Institute of Medical Sciences (AIIMS), New Delhi, India in collaboration with Institute of Genomics and Integrative Biology (IGIB), New Delhi, India. From October 2017 to March 2020, we included consecutive stroke patients aged 18 years and above, ischemic or hemorrhagic confirmed by neuroimaging and clinical diagnosis admitted within 24 hours of symptom onset to the neurology wards and/or emergency department of AIIMS, New Delhi. All included patients had clinical signs consistent with the definition of stroke given by the American Heart Association (AHA)/American Stroke Association (ASA).^11^ We also included a group of stroke mimics presenting within 24 hours with stroke-like symptoms. We excluded patients with a previous history of stroke, admission after the 24-hour onset of the qualifying event, cerebral vein thrombosis, chronic liver and/or kidney disease, documented history of cancer, pregnancy, and unwilling to provide written informed consent.

The study was approved by the Local Institutional Ethics Committee of AIIMS, New Delhi (Ref. No. IECPG-395/28.09.2017). We obtained written informed consent from all the recruited patients or legally authorized representative prior to collecting blood samples and clinical history. The study was divided into two phases: (1) discovery phase and (2) validation phase. The study flow diagram is represented in Figure 1.

**Figure 1:**
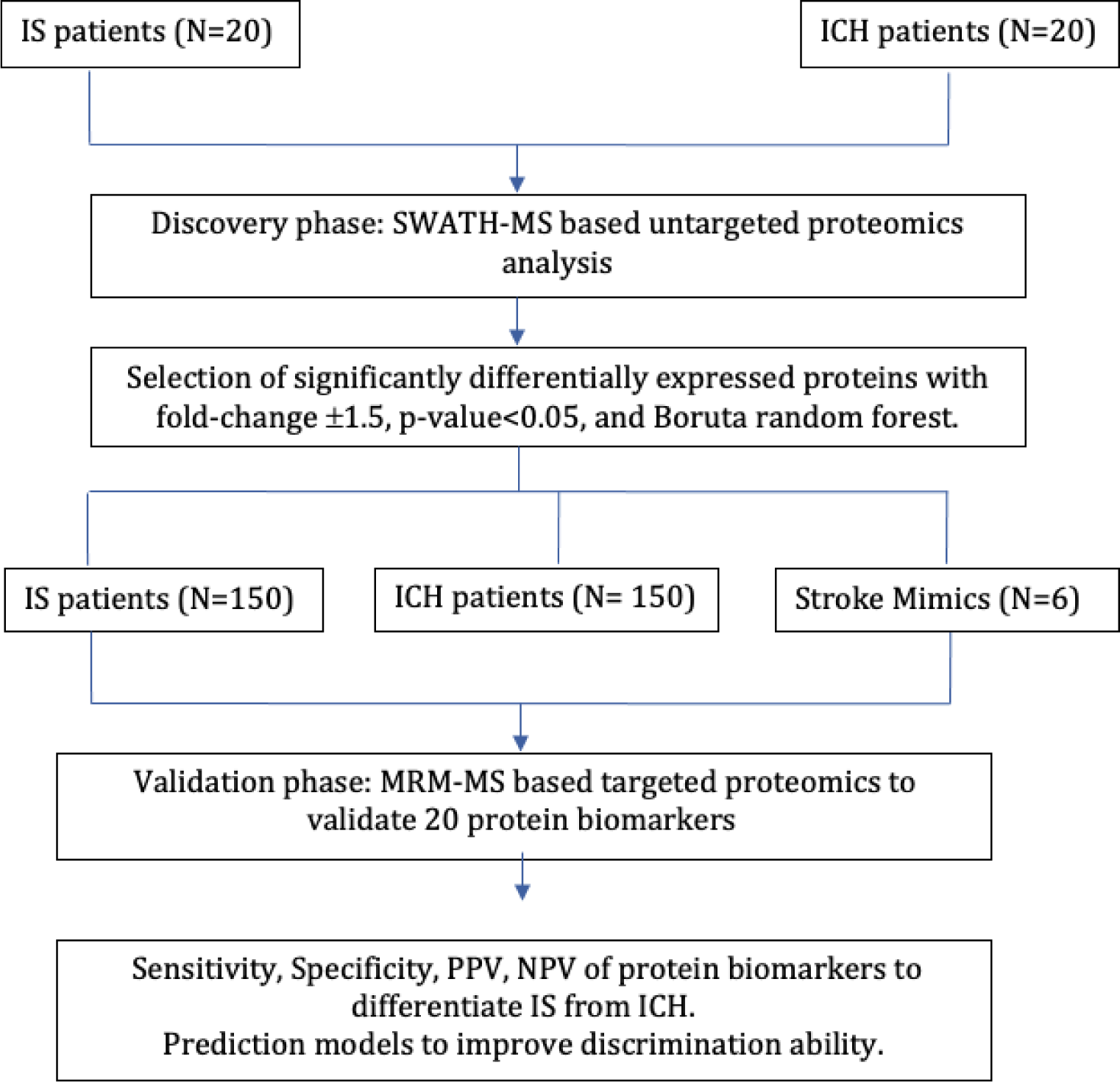
Study flow diagram

### Sample size

For the discovery phase, based on the feasibility, budget, and time-frame of the study, we selected 40 stroke patients comprising of 20 IS and 20 ICH. The literature suggests a sample size of 10 to 30 for conducting a pilot/discovery phase study.^12,13^

For the validation phase, to achieve a sensitivity and specificity of 90% with 5% margin of error and 5% drop out rate, we estimated a final sample size of 300 comprising 150 IS and 150 ICH patients^14^ using STATA version 13.0 software.

### Blood sample collection

We collected five ml of peripheral blood samples in serum vacutainer tubes from stroke patients and mimics within 24-hour onset. For serum collection, it was left standing at room temperature for 30 minutes until clotted. It was then centrifuged at 3000g for 10 minutes, after which the serum was separated into cryovials. Five aliquots of each sample (100µl) were prepared and stored at -80°C until further analysis.

### Sample preparation, Reduction, alkylation, and trypsin digestion

Refer to previously published paper.^15^

### DISCOVERY PHASE

#### SWATH-MS data acquisition

Refer to previously published paper.^15^

#### Bioinformatic analysis

We used a high-pH fractionated peptide library for human serum proteins (obtained from SCIEX) comprising 465 proteins. SWATH peaks were extracted using this library in SWATH 2.0 microapp in PeakView 2.2 software (SCIEX), excluding shared peptides. The processing settings for peak extraction were: maximum 10 peptides per protein, 5 transitions per peptide, >95% peptide confidence threshold, 1% peptide false discovery rate (FDR). XIC extraction window was set to 55 minutes with 75 ppm XIC Width. Data were normalized using total area sum normalization and log^2^ transformed. Batch correction was performed for removing the non-biological experimental variations and was visualized using the Principal Component Analysis (PCA) plots. Differentially expressed proteins were selected using two criteria: (i) p-value <0.05 and ± 1.5 fold change cut-offs (>1.5 for upregulated and <0.67 for downregulated proteins); or (ii) confirmed/tentative selection in the Boruta random forest feature selection method. The STRING 11 online tool^16^ was used to create the protein network of the differentially expressed proteins between various conditions. Further, protein-protein interaction network and the functional enrichment analyses were conducted using Cytoscape 3.9.0 software.^17^

## VALIDATION PHASE

Differentially expressed proteins obtained in the discovery phase and five proteins identified in our meta-analysis^9^ were validated using targeted proteomics in the validation phase.

### Peptide selection for Multiple Reaction Monitoring (MRM)-based targeted proteomics

Peptide selection was performed using search results from ProteinPilot,^18^ PeptideAtlas^19^ or in-silico generated peptides of proteins using Expasy PeptideCutter tool.^20^ Peptides with +2 and +3 charges were considered for MRM and for each peptide, 5-6 fragment ions were used for identification. Peptide selection for the list of shortlisted proteins is given in supplementary Table 1.

### MRM data acquisition

Tryptic peptides obtained after digestion were desalted using reversed-phase Oasis HLB cartridges (Waters, Milford, MA) as per manufacturer’s protocol. The peptide mixture was dried using a vacuum centrifuge, and the peptides were resuspended in 0.1% formic acid at a final concentration of 1μg/μl. A heavy labeled peptide for Apo A1 (QGLLPVLESF**K; K**=Lysine-13C6,15N2) protein was spiked-in the resolubilized plasma digest at a final concentration of 1ng/μl.

The targeted MRM-MS^21^ analysis of the tryptic peptides was performed on a TSQ Altis (Thermo Fisher, San Jose, CA). The instrument was equipped with an H-ESI ion source. A spray voltage of 3.5 keV was used with a heated ion transfer tube set at a temperature of 325°C. Chromatographic separations of peptides were performed on Vanquish UHPLC system (Thermo Fisher, San Jose, CA).

Ten μl sample was injected and peptides were loaded on an ACQUITY UPLC BEH C18 column (130Å, 1.7 µm, 2.1 mm X 100 mm, Waters) from a cooled (4°C) autosampler and separated with a linear gradient of water (buffer A) and acetonitrile (buffer B), containing 0.1% formic acid, at a flow rate of 300µl/minute in 30 minutes gradient run.

### Statistical analysis

Skyline version 21.1 was used to analyze the MRM data.^22^ The area for each peptide was spike-in normalized using a heavy labeled peptide for Apo A1 protein and then log^2^ transformed. The random forest-based imputation was performed for peptides with <10% missing values.^23^ The normality of the data was assessed using the Shapiro-Wilk test.

The results of our study were reported as per the Standard for Reporting Diagnostic Accuracy (STARD) guidelines.^24^ For each comparison, IS was taken as the endpoint and univariable logistic regression analysis was conducted using odds ratio (OR) and 95% confidence interval (CI). Receiver operating characteristic (ROC) curve analyses were performed for biomarkers differentiating IS from ICH and stroke mimics. The optimal cut-off points were determined using the Youden Index. The sensitivity, specificity, positive predictive value (PPV), and negative predictive value (NPV) of each biomarker were determined. Since the aim of this study was to detect and diagnose IS, cut-offs for each biomarker were selected having the highest sensitivity values to detect IS and to rule out ICH or mimics with the best available specificity values.

Prediction models were developed to determine the accuracy and precision of protein biomarkers and their discrimination ability was evaluated using area under the curve (AUC)/c-statistic. The clinical variables with p-value <0.1 in the univariable analysis along with demographic variables like age and sex, were included in a backward stepwise multivariable logistic regression analysis. Afterward, biomarkers were added incrementally to this model. Those biomarkers that contributed to at least a 1% increase in the AUC/c-statistic were retained in the model. Another prediction model was developed to determine the diagnostic potential of biomarkers alone in differentiating IS. For each prediction model, the degree of multicollinearity was tested using the variance inflation factor (VIF) and predictors with VIF value >2.5 were removed from the final model. The discrimination ability (AUC) between various prediction models was compared and quantitatively assessed using the Hanley-McNeil test p-value.^25^ At different pre-test probabilities of the disease, the corresponding post-test probability, and the multi-level likelihood ratio were determined. The integrated discrimination improvement (IDI) index was calculated to assess the added value of the biomarkers to the clinical predictive models.^26,27^ The goodness-of-fit between the two hierarchical prediction models was evaluated using the Likelihood Ratio (LR) test based on the ratio of their likelihoods.^28^ The selection of the best prediction model was done using Akaike’s Information Criteria (AIC)^29^ and Bayesian Information Criteria (BIC).^30^ The statistical analyses were conducted in R version 3.6.2 and STATA version 13.1.

### Data availability

The mass spectrometry proteomics data have been deposited to the ProteomeXchange Consortium via the PRIDE^31^ partner repository with the dataset identifier PXD032917.

## Results

### Results from the Discovery Phase

We enrolled 40 patients in the discovery phase: 20 IS and 20 ICH patients recruited within 24 hours. Between IS and ICH cases, there was no significant difference in the mean age (p=0.12) and mean blood sampling time from symptom onset (p=0.86). The baseline characteristics of the patients are given in supplementary Table 2.

Serum proteomic profiles were compared between 20 IS and 20 ICH cases using the SWATH-MS approach. We quantified 375 proteins at 1% peptide FDR between IS and ICH. The total ion chromatogram and the PCA plots depicting batch variations are given in supplementary figure 1 and supplementary figure 2.

We identified 14 differentially expressed proteins using the fold change and p-value criteria. Three proteins were upregulated, while 11 were downregulated in IS compared to ICH (Figure 2a). We further identified 11 confirmed/tentative features between IS and ICH using the Boruta random forest method (Figure 2b and supplementary Table 3). Five proteins (UniProt IDs: P01023, P01833, P01764, P19652, and P02654) were common using both protein selection criteria. Therefore, after combining the distinctly expressed proteins using both criteria, 20 differentially expressed proteins were identified between IS and ICH within 24 hours (Table 1). A heatmap of these 20 proteins is depicted in Figure 2c.

**Figure 2a:**
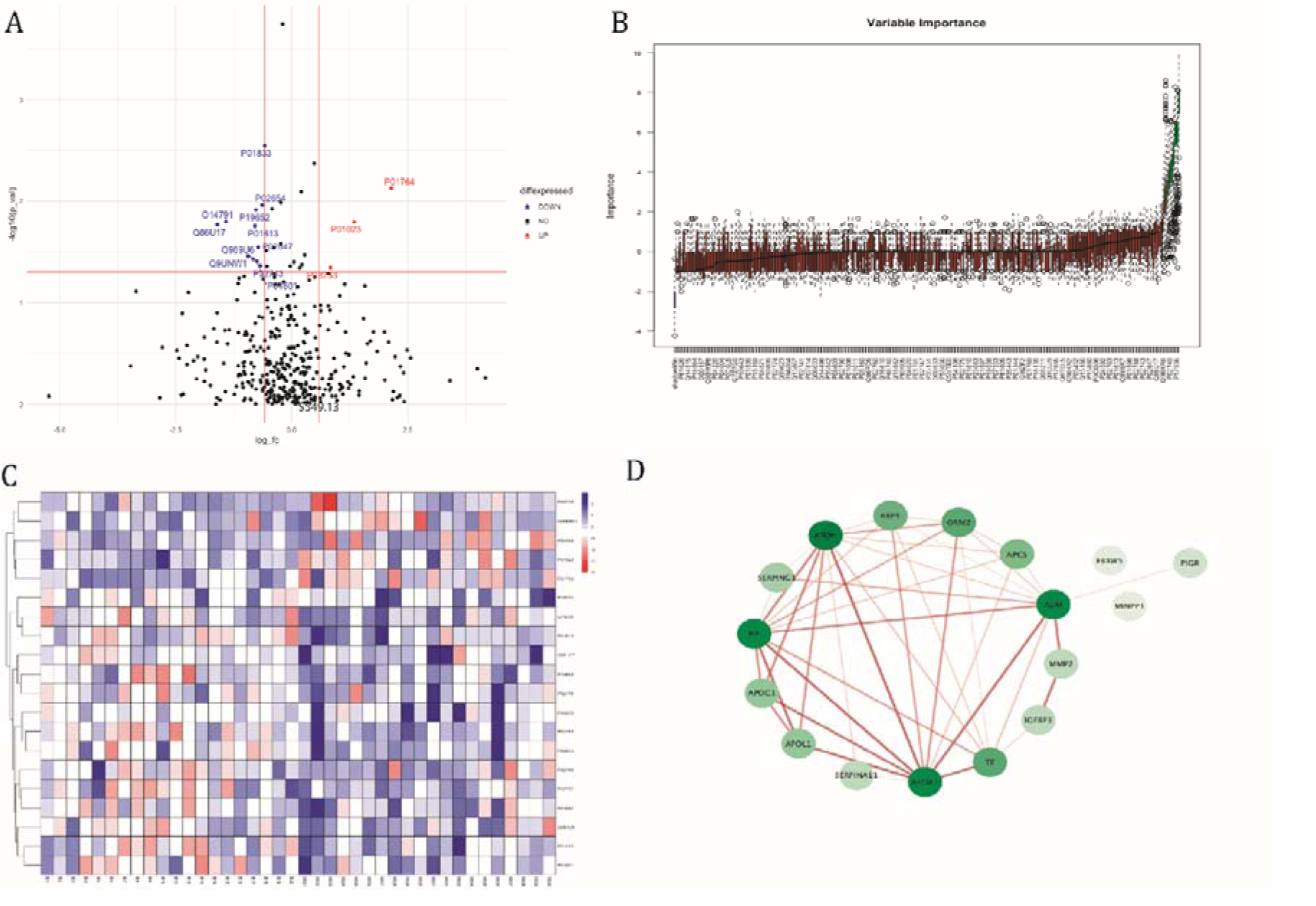
Volcano plot depicting the log^2^ fold change on the x-axis and -log^10^ p-value on the y-axis for the upregulated and downregulated proteins in 20 IS cases compared to 20 ICH cases within 24 hours. Figure 2b: Feature selection using the Boruta random forest between 20 IS and 20 ICH cases within 24 hours. Figure 2c: Heatmap showing the distribution and expression pattern of 20 significantly differentially expressed proteins between IS and ICH. Figure 2d: Protein-protein interaction network analysis of differentially expressed proteins between IS and ICH using the Cytoscape software. The colour of nodes and edges represents the degree of interaction and interaction score.

**Table 1:** List of significantly differentially expressed proteins between IS and ICH within 24 hours of symptom onset using Fold change with p-value and Boruta random forest feature selection criteria

| S. No | UniProt ID | Protein Name (Gene annotation) | Fold Change* | P-value | Boruta decision |
| --- | --- | --- | --- | --- | --- |
| 1 | P01764 | Ig heavy chain V-III region VH26 | **4.42** | **0.007** | **Confirmed** |
| 2 | P01023 | Alpha-2-macroglobulin (GN=A2M) | **2.54** | **0.02** | **Confirmed** |
| 3 | P08253 | 72 kDa type IV collagenase (GN=MMP2) | **1.78** | **0.04** | Rejected |
| 4 | P05155 | Plasma protease C1 inhibitor (GN=SERPING1) | 1.12 | **0.04** | **Confirmed** |
| 5 | P00738 | Haptoglobin (GN=HP) | 0.97 | **0.04** | **Tentative** |
| 6 | P02749 | Beta-2-glycoprotein 1 (GN=APOH) | 0.95 | 0.17 | **Confirmed** |
| 7 | P02753 | Retinol-binding protein 4 (GN=RBP4) | 0.87 | **0.0001** | **Confirmed** |
| 8 | P02787 | Serotransferrin (GN=TF) | 0.82 | 0.07 | **Confirmed** |
| 9 | P01833 | Polymeric immunoglobulin receptor (GN=PIGR) | **0.66** | **0.003** | **Confirmed** |
| 10 | P02654 | Apolipoprotein C-I OS=Homo sapiens (GN=APOC1) | **0.64** | **0.01** | **Confirmed** |
| 11 | P02743 | Serum amyloid P-component (GN=APCS) | **0.62** | **0.04** | Rejected |
| 12 | P02647 | Apolipoprotein A-I (GN=APOA1) | **0.61** | **0.03** | Rejected |
| 13 | P01601 | Ig kappa chain V-I region HK101 (Fragment) | **0.59** | **0.04** | Rejected |
| 14 | P19652 | Alpha-1-acid glycoprotein 2 (GN=ORM2) | **0.59** | **0.01** | **Tentative** |
| 15 | P01613 | Ig kappa chain V-I region Ni | **0.58** | **0.02** | Rejected |
| 16 | Q969U6 | F-box/WD repeat-containing protein 5 (GN=FBXW5) | **0.56** | **0.04** | Rejected |
| 17 | Q9UNW1 | Multiple inositol polyphosphate phosphatase 1 (GN=MINPP1) | **0.52** | **0.03** | Rejected |
| 18 | O14791 | Apolipoprotein L1 (GN=APOL1) | **0.37** | **0.01** | Rejected |
| 19 | Q86U17 | Serpin A11 (GN=SERPINA11) | **0.33** | **0.02** | Rejected |
| 20 | P17936 | Insulin-like growth factor-binding protein 3 (GN=IGFBP3) | **0.21** | 0.08 | **Confirmed** |

\*Fold change is a ratio representing the change of protein concentration between IS and ICH patients.
**Bold values:** Fold change >1.5 or <0.67, p-value<0.05 and confirmed/tentative in Boruta random forest.
Abbreviations: A2M- Alpha-2-macroglobulin, MMP2- Matrix metalloproteinase 2 (72 kDa type IV collagenase), SERPING1- Plasma protease C1 inhibitor, HP- Haptoglobin, APOH- Apolipoprotein H (Beta-2-glycoprotein 1), RBP4- Retinol Binding protein 4, TF- Serotransferrin, PIGR- Polymeric immunoglobulin receptor, APOC1- Apolipoprotein C-I, APCS- Serum amyloid P-component, APOA1- Apolipoprotein A1, ORM2- Orosomucoid (Alpha-1-acid glycoprotein 2), FBXW5- F-
box/WD repeat-containing protein 5, MINPP1- Multiple inositol polyphosphate phosphatase 1, APOL1- Apolipoprotein L1, SERPINA11- Serpin A11, IGFBP3- Insulin-like growth factor-binding protein 3.

Interaction network analysis observed that 15 proteins formed a highly connected network showing molecular interactions with other proteins (Figure 2d). APOH had the highest degree of interaction with other proteins followed by APOA1, A2M, and HP. In our network, 10 protein-protein interactions had an interaction score of >0.90. Functional enrichment analysis identified molecular pathways/processes involving blood microparticles, extracellular space, lipoprotein particles, cholesterol metabolism, platelet degranulation, and post translational protein phosphorylation that were significantly different between IS and ICH (supplementary Table 4).

### Results from the Validation Phase

We included 306 cases in the validation phase; 300 stroke patients (150 IS and 150 ICH) and 6 stroke mimics within 24 hours. The mean age of IS, ICH, and stroke mimics were 54.59±15.54, 55.30±12.72, and 47.83±18.25 years, respectively. IS, ICH, and stroke mimics consisted of 64.67%, 65.33%, and 66.67% males. The mean blood sampling time (in hours) from symptom onset was 15.29±5.55 in IS, 14.97±6.55 in ICH, and 8.75±5.07 in stroke mimics. The 6 stroke mimics included patients suffering from hypoglycaemia (2), vertigo (2), syncope (1), and seizure (1) (supplementary Table 5 and supplementary Table 6).

We validated 20 differentially expressed proteins identified in the discovery phase and five proteins (GFAP, BNP, D-dimer, MMP-9, and UCH-L1) identified in our published meta-analysis.^9^ While selecting the peptides of interest for MRM, unique peptides for two proteins (UniProt IDs: P01764 and P01601) could not be determined, while the unique peptide (TDISMSDFENSR) for PIGR protein (UniProt ID: P01833) was not detectable in any sample, thus they were excluded from the final analysis. Therefore, 22 proteins (17 from discovery phase and five from meta-analysis) were selected in our validation phase. One unique peptide per protein was used for quantification. The final list of unique peptides and quantifier fragment ion for each protein is given in supplementary Table 7.

### Univariable analysis in the Validation Phase

In the univariable analysis, the protein concentration of 15 out of 22 biomarkers were different between IS and ICH (p-value <0.1) (supplementary Table 8). The diagnostic characteristics for the 15 categorized biomarkers are given in Table 2. In a subgroup analysis conducted within 6 hours, nine of 22 biomarkers significantly differentiated 12 IS from 26 ICH cases (supplementary Table 9 and supplementary Table 10). In a secondary analysis, only three biomarkers differentiated IS from stroke mimics within 24 hours (supplementary Table 11).

**Table 2:** The diagnostic potential of protein biomarkers assessed in the validation phase of the study to differentiate ischemic stroke from intracerebral hemorrhage within 24 hours of onset

| S. No | Protein Biomarker (UniProt ID) | OR (95% CI) | p-value | Cutoff | Sensitivity (95% CI) | Specificity (95% CI) | PPV (95% CI) (For IS) | NPV (95% CI) (For ICH) |
| --- | --- | --- | --- | --- | --- | --- | --- | --- |
| 1 | GFAP (P14136) | 0.06 (0.03-0.12) | <0.001 | <16.54 | 93% (88-97%) | 55% (46-63%) | 67% (60-74%) | 89% (81-95%) |
| 2 | MMP-9 (P14780) | 0.15 (0.07-0.32) | <0.001 | <16.52 | 94% (89-97%) | 30% (23-38%) | 57% (51-63%) | 83% (71-92%) |
| 3 | UCH-L1 (P09936) | 0.24 (0.12-0.44) | <0.001 | <14.03 | 90% (84-94%) | 32% (25-40%) | 57% (50-63%) | 76% (64-86%) |
| 4 | MINPP1 (Q9UNW1) | 0.39 (0.24-0.62) | <0.001 | <19.93 | 73% (65-80%) | 49% (41-58%) | 59% (51-66%) | 64% (55-73%) |
| 5 | APO-A1 (P02647) | 0.13 (0.07-0.26) | <0.001 | <21.92 | 92% (86-96%) | 39% (31-48%) | 60% (54-67%) | 83% (72-91%) |
| 6 | Alpha-1-acid-Glycoprotein (ORM2) (P19652) | 0.33 (0.17-0.63) | 0.001 | <20.29 | 24% (17-32%) | 91% (85-95%) | 72% (57-84%) | 54% (48-61%) |
| 7 | BNP (N=179) (P16860) | 0.38 (0.20-0.72) | 0.003 | <9.46 | 74% (64-83%) | 47% (36-58%) | 59% (49-68%) | 65% (52-76) |
| 8 | Beta-2-glycoprotein 1 (APOH) (P02749) | 0.25 (0.10-0.65) | 0.004 | <20.25 | 96% (91-98%) | 14% (9-21%) | 53% (47-59%) | 78% (58-91%) |
| 9 | RBP4 (P02753) | 0.49 (0.30-0.80) | 0.005 | <17.32 | 75% (68-82%) | 40% (32-48%) | 56% (48-63%) | 62% (51-71%) |
| 10 | MMP-2 (P08253) | 0.54 (0.33-0.89) | 0.01 | <18.63 | 40% (32-48%) | 73% (65-80%) | 60% (48-70%) | 55% (48-62%) |
| 11 | APO-C1 (P02654) | 1.94 (1.15-3.29) | 0.01 | >16.26 | 80% (73-86%) | 33% (25-41%) | 54% (47-61%) | 62% (50-73%) |
| 12 | Alpha-2-Macroglobulin (P01023) | 0.56 (0.34-0.93) | 0.02 | <18.70 | 75% (68-82%) | 37% (29-45%) | 54% (47-61%) | 60% (49-70%) |
| 13 | IGFBP3 (P17936) | 0.53 (0.30-0.95) | 0.03 | <17.10 | 85% (78-90%) | 25% (18-33%) | 53% (46-59%) | 62% (49-74%) |
| 14 | Haptoglobin (P00738) | 1.81 (1.01-3.22) | 0.04 | >17.21 | 85% (78-90%) | 25% (18-32%) | 53% (46-59%) | 62% (48-74%) |
| 15 | Serotransferrin (P02787) | 0.51 (0.26-0.99) | 0.04 | <21.02 | 90% (84-94%) | 18% (12-25%) | 52% (46-58%) | 64% (48-78%) |

The cut-off values represent the Log<sub>2</sub> normalized protein concentrations.
**Abbreviations:** OR: Odds Ratio; CI: Confidence Interval; PPV: Positive Predictive Value; NPV: Negative Predictive Value; APO- Apolipoprotein; APOH: Apolipoprotein H (Beta-2-glycoprotein 1); MINPP1: Multiple inositol polyphosphate phosphatase 1; ORM2: Orosomucoid 2 (Alpha-1-acid glycoprotein 2); RBP4: Retinol Binding Protein 4; MMP- Matrix Metalloproteinase; MMP2: Matrix Metalloproteinase 2 (72 kDa type IV collagenase); A2M: Alpha-2-Macroglobulin; IGFBP3: Insulin-like growth factor-binding protein 3; FBXW5: F-box/WD repeat-containing protein 5; APCS: Serum amyloid P-component; SERPING1: Serpin Family G Member 1 (Plasma protease C1 inhibitor); GFAP: Glial Fibrillary Acidic Protein; UCH-L1: Ubiquitin C-Terminal Hydrolase L1; BNP: Brain Natriuretic Peptide.

### Multivariable regression analysis in the validation phase

#### Clinical model

Five clinical variables including sex (OR 2.12; 95%CI 1.12-4.02), hypertension (OR 0.40; 95%CI 0.23-0.70), atrial fibrillation (OR 9.13; 95%CI 2.01-41.57), current smoking (OR 3.19; 95%CI 1.70-5.96), and NIHSS score at admission (OR 0.87; 95%CI 0.84-0.90) were independent predictors of IS (AUC/c-statistic of 81%) (Table 3).

**Table 3:** Multivariable prediction models to differentiate IS from ICH within 24 hours

| S. No | Predictor variables | OR (95% CI) | P-value |
| --- | --- | --- | --- |
| **Clinical model discrimination ability (AUC)/ c-statistic: 0.81 (0.76-0.86)** |  |  |  |
| 1. | Sex | 2.12 (1.12-4.02) | 0.02 |
| 2. | Hypertension | 0.40 (0.23-0.70) | 0.001 |
| 3. | Atrial Fibrillation | 9.13 (2.01-41.57) | 0.004 |
| 4. | Current Smoking | 3.19 (1.70-5.96) | <0.001 |
| 5. | NIHSS score at admission | 0.87 (0.84-0.90) | <0.001 |
| **Biomarker model discrimination ability (AUC)/ c-statistic: 0.88 (0.84-0.92)** |  |  |  |
| 1. | GFAP (<16.54) | 0.06 (0.03-0.16) | <0.001 |
| 2. | MMP9 (<16.52) | 0.16 (0.06-0.42) | <0.001 |
| 3. | APO-C1 (>16.26) | 6.56 (3.20-13.45) | <0.001 |
| 4. | Haptoglobin (>17.21) | 2.88 (1.26-6.58) | 0.01 |
| 5. | APO-A1 (<21.92) | 0.28 (0.09-0.81) | 0.02 |
| 6. | UCH-L1 (<14.03) | 0.35 (0.14-0.84) | 0.02 |
| 7. | Serotransferrin (<21.02) | 4.40 (1.30-14.85) | 0.02 |
| 8. | ORM2 (<20.29) | 0.38 (0.15-0.95) | 0.04 |
| **Combined model discrimination ability (AUC)/ c-statistic: 0.92 (0.89-0.95)** |  |  |  |
| 1. | Sex | 1.82 (0.83-3.99) | 0.13 |
| 2. | Hypertension | 0.38 (0.19-0.76) | 0.007 |
| 3. | Atrial Fibrillation | 21.92 (3.17-151.35) | 0.002 |
| 4. | Current Smoking | 3.00 (1.37-6.55) | 0.006 |
| 5. | NIHSS score at admission | 0.87 (0.84-0.91) | <0.001 |
| 6. | GFAP (<16.54) | 0.04 (0.02-0.11) | <0.001 |
| 7. | MMP9 (<16.52) | 0.09 (0.03-0.28) | <0.001 |
| 8. | APO-C1 (>16.26) | 5.76 (2.66-12.47) | <0.001 |

**Abbreviations:** OR: Odds Ratio; CI: Confidence Interval; AUC: Area Under the Curve; NIHSS: National Institutes of Health Stroke Scale; GFAP: Glial Fibrillary Acidic Protein; MMP9: Matrix Metalloproteinase 9; APO: Apolipoprotein; ORM2: Orosomucoid 2 (Alpha-1-acid glycoprotein 2); UCH-L1: Ubiquitin C-Terminal Hydrolase L1.
**Clinical model:** Adjusted for age, sex, time taken to reach the hospital, hypertension, diabetes, atrial fibrillation, current smoking, mild alcohol consumption, family history of diabetes, family history of heart attack, NIHSS score at admission and TLC. **Biomarker model:** Adjusted for GFAP, UCH-L1, MMP2, MMP9, RBP4, MINPP1, APO-A1, APO-C1, APOH, ORM2, Haptoglobin, Serotransferrin, A2M, and IGFBP3. **Combined model:** Adjusted for sex, hypertension, atrial fibrillation, current smoking, NIHSS score at admission, GFAP, MMP9, and APO-C1.

#### Biomarker model

Eight protein biomarkers including GFAP (OR 0.06; 95%CI 0.03-0.16), MMP-9 (OR 0.16; 95%CI 0.06-0.42), APO-C1 (OR 6.56; 95%CI 3.20-13.45), APO-A1 (OR 0.28; 95%CI 0.09-0.81), Haptoglobin (OR 2.88; 95%CI 1.26-6.58), UCH-L1 (OR 0.35; 95%CI 0.14-0.84), ORM2 (OR 0.38; 95%CI 0.15-0.95), and Serotransferrin (OR 4.40; 95%CI 1.30-14.85) were independent predictors of IS (AUC/c-statistic of 88%) (Table 3).

#### Combined model (clinical variables + protein biomarkers)

Proteins that were significant in the biomarker model were incrementally added to the clinical model. In the combined prediction model, four clinical variables including hypertension (OR 0.38; 95%CI 0.19-0.76), atrial fibrillation (OR 21.92; 95%CI 3.17-151.35), current smoking (OR 3.00; 95%CI 1.37-6.55), and NIHSS score at admission (OR 0.87; 95%CI 0.84-0.91), and three protein biomarkers including GFAP (OR 0.04; 95%CI 0.02-0.11), MMP-9 (OR 0.09; 95%CI 0.03-0.28), and APO-C1 (OR 5.76; 95%CI 2.66-12.47) were independent predictors of IS (AUC/c-statistic of 92%) (Table 3).

#### Comparing the diagnostic performance of prediction models

We compared the ROC curves of three prediction models using the Hanley and McNeil test. We observed that the combined model had significantly higher AUC compared to biomarker model (p=0.036) and clinical model (p<0.0001) (Figure 3).

**Figure 3:**
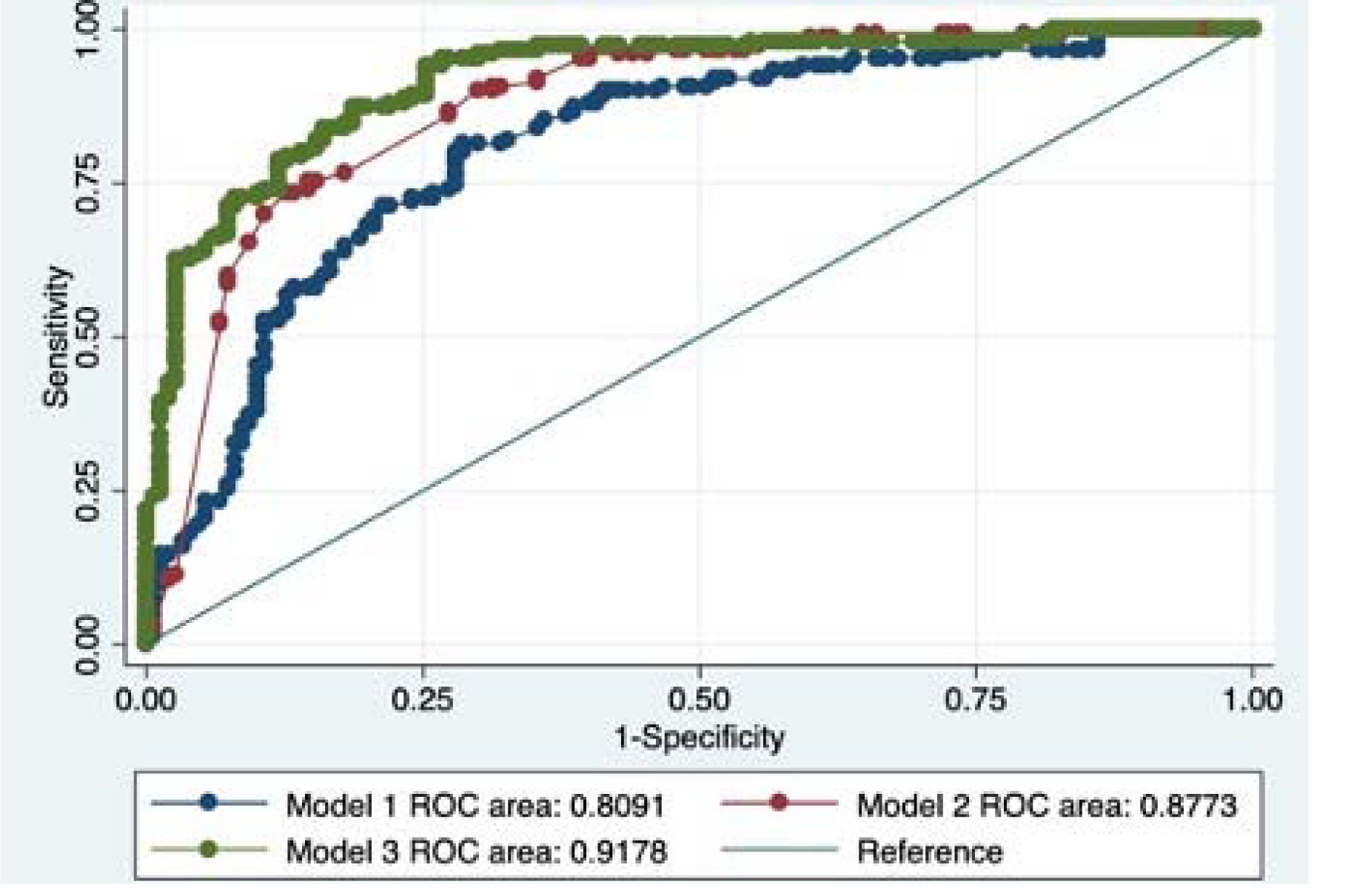
A comparison between the ROC curves of Clinical prediction model (blue curve), Biomarker prediction model (red curve), and Combined prediction model (green curve) to differentiate IS from ICH within 24 hours.

The IDI index suggested that combined model increased the discrimination ability by 13% (p<0.001) compared to biomarker model and by 26% (p<0.001) compared to clinical model.

The goodness-of-fit between the clinical and combined models (hierarchical models) was compared using the likelihood ratio test. The addition of three protein biomarkers (GFAP, MMP-9, and APO-C1) to the clinical variables significantly increased the likelihood ratio by 102.74 (p<0.001). Further, combined model including clinical and biomarker variables, had the least AIC and BIC values than clinical model.

#### Comparison between prediction models at different pre-test probabilities

Combined model including clinical variables and protein biomarkers (GFAP, MMP-9, APO-C1), improved the pre-test probability of diagnosing an IS from 30%, 50%, and 70% to a post-test probability of 57%, 82%, and 95%, respectively. Combined model had the best sensitivity, specificity, and likelihood ratios at all the three pre-test probability cut-offs compared to clinical and biomarker models. At a pre-test probability of 30%, combined model had the highest NPV of 94% to rule out ICH (Table 4).

**Table 4:**
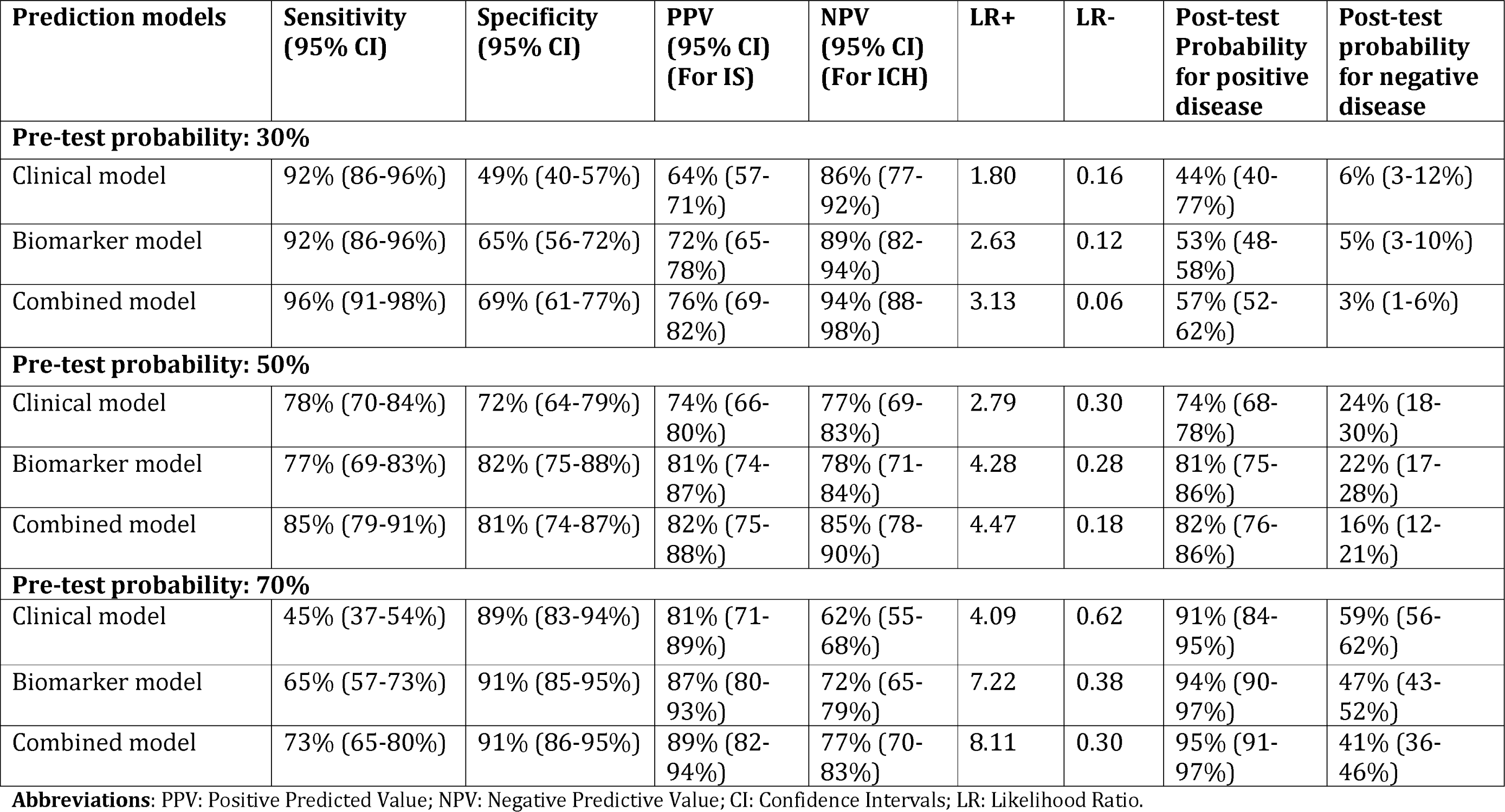
Comparison in the diagnostic performance of the three prediction models at different pre-test probabilities

### Discussion

In the discovery phase, our study identified 20 differentially expressed proteins between IS and ICH. Of these 20 proteins, in the validation phase, we identified three biomarkers (GFAP, MMP9, APOC1) that independently differentiated IS from ICH in multivariable analyses. When these three biomarkers were added to the prediction models containing clinical variables, they significantly improved the discrimination ability to detect IS. GFAP and MMP9 further differentiated the two stroke subtypes in a subgroup analysis within 6 hours. In a secondary analysis, we identified three biomarkers (APO-L1, BNP, FBXW5) that differentiated IS from stroke mimics. To the best of our knowledge, this is the first study that has utilized the discovery-based label-free SWATH-MS proteomics and targeted MRM proteomics to identify and validate differentially expressed protein biomarkers in the blood of patients with IS and ICH within 24 hours of symptom onset.

Our findings highlight the importance of a panel-based approach in diagnostic biomarker research in stroke. Our study observed that a combined panel of biomarkers and clinical variables detected IS from ICH with an accuracy of 92%. Only a few studies in the past have adopted the panel-based approach. The Stroke-Chip study in 2017^32^ included a panel of clinical variables and biomarkers in which only NT-proBNP independently differentiated IS from ICH with an accuracy of 75.7%. Earlier, Montaner et al. in 2012^33^ observed that in a combined panel, S100B and RAGE independently differentiated IS from ICH with an improved accuracy of 84%. Thus, future studies must focus on testing the diagnostic potential of their biomarkers in multivariable prediction models to identify independent biomarkers for IS diagnosis.

For the first time our study used a label-free proteomics approach to detect the levels of proteins identified in the literature such as GFAP, MMP9, BNP, D-dimer, and UCH-L1 in stroke patients. We focussed on finding a combination of biomarkers with/without clinical variables which can detect IS with high sensitivity and a high NPV to rule out ICH. The poor specificity values observed in most of the biomarkers when analyzed alone, were improved significantly when combined in a panel.

Our network analysis identified an interaction network of 15 proteins that was highly connected. The most common significant pathways underlying the proteins differentiating IS from ICH included cholesterol metabolism and lipid-related processes, platelet degranulation, and pathways related to extracellular space and matrix.

Thus far, no proteomics study conducted in stroke has identified biomarkers within the 24-hour time window. In a recent study, Malicek et al. 2021^34^ used a similar discovery-based label-free proteomics approach and identified nine proteins in a small sample of 3 IS and 4 ICH patients. They collected blood samples between 1 to 15 days and lacked a validation phase. Of these nine proteins, serum amyloid P-component was also differentially expressed in our discover phase. Another study by Zhang et al. 2021^35^ collected serum within five days and identified two metabolic biomarkers between IS and ICH using targeted metabolomics. Lopez et al. (2012)^36^ used targeted proteomics and identified a combination of APO-CIII and APO-AI that differentiated IS from ICH with an AUC of 0.92. In our discovery phase, we also observed that APO-AI was significantly downregulated in IS by 0.61-fold compared to ICH.

Several studies in the past have used immunoassays to identify candidate protein biomarkers for differentiating IS from ICH. GFAP is the most extensively studied protein and several systematic reviews and meta-analyses,^37–40^ including those published by our group^8,9,41^ also reiterate our findings that GFAP efficiently differentiates the two stroke subtypes with high sensitivity and/or specificity within 24 as well as 6 hours. With regards to MMP9, our findings are in line with several other published studies^33,42–44^ and meta-analysis,^9^ that observed that MMP-9 has significantly lower levels in IS than ICH and efficiently differentiates the two stroke subtypes within 24 hours.

Previous studies have had contradictory results regarding the APO-C1 levels in IS and ICH patients. In our study, we detected higher levels of APO-C1 in IS than ICH. The increased levels of APO-C1 during acute IS might be related to its key role in lipid metabolism, also observed in the pathway analysis in our study. One study by Allard et al. (2004)^45^ corroborates our findings as they observed higher levels of APO-C1 in IS than ICH. However, a study by Walsh et al. (2016)^46^ observed that in a small sample of 14 IS and 23 ICH cases, the median APO-CI levels were slightly lower in IS compared to ICH. Therefore, more adequately powered studies are required to confirm the levels of APO-C1 in stroke subtypes.

### Strengths of our study

This study provides crucial insights into the pathophysiology of IS and ICH in acute stages. We validated the diagnostic potential of discovered biomarkers in a large cohort of patients using a high-throughput proteomics approach. Our study was conducted in resource-limited settings wherein there is an urgent need for an alternative approach toward rapid stroke diagnosis. Our study analyzed the individual protein profile of each patient using proteomics in both discovery and validation phases without pooling any sample. Since, pooled samples do not reflect the diseased/non-diseased condition of a single person and may produce false-positive or false-negative results.

### Limitations of the study

Certain inherent limitations associated with our study must be kept in mind when interpreting its results. Firstly, the serum samples were obtained only from the Indian population; thus, further studies conducted in ethnically diverse populations are required to validate the generalizability of the biomarkers identified in our study. Secondly, relative quantification of proteins was done in the validation phase. To determine the biomarker levels with absolute quantification, future studies must be conducted on these biomarkers using either labelled peptide-based targeted proteomics or standard immunoassays. Lastly, the number of stroke mimics included in this study were very less, therefore, findings were only exploratory.

### Future directions

The literature on proteomics studies for diagnostic biomarkers in stroke is mainly limited to the discovery phase, while the validation phase is lacking. Therefore, studies must look to validate their discovered biomarkers in a large independent cohort using either standard immunoassays or targeted proteomics approaches using MRM. Since stroke is a multifactorial disorder, a single biomarker approach might not be appropriate, and instead studies must look for a panel of biomarkers that may increase the overall sensitivity and specificity of the diagnosis. The ability of the biomarkers to independently diagnose IS must be evaluated in multivariable regression models consisting of potential clinical predictors along with independent protein biomarkers. Studies should also examine the improvement in the discrimination ability and the incremental value of any biomarker over clinical variables using various statistical approaches. Future studies should utilize the advanced label-free proteomics technology for biomarker research in stroke. They require a minimal sample volume at a low cost per sample. This approach would be convenient for studies investigating a panel of biomarkers. Studies must avoid pooling of samples when conducting their proteomics experiments to avoid false positive/false negative results.

### Conclusion

Our study identified 20 differentially expressed proteins between IS and ICH in the discovery phase using SWATH-MS proteomics with a strong interaction network for 15 proteins. The validation phase highlighted the potential of 15 biomarker candidates in differentiating IS from ICH within 24 hours. The prediction model including three protein biomarkers (GFAP, MMP9, and APOC1) and clinical variables, had the best discrimination ability and increased the post-test probability of detecting IS with high sensitivity, specificity, and likelihood ratios. Protein biomarkers along with clinical predictors significantly improved the discrimination ability of the model in differentiating IS from ICH within 24 hours. Our results must be validated in other populations using absolute quantification to confirm the generalizability of our findings and the diagnostic performance of the identified biomarkers.

### Contributors

DV conceptualized the idea of this research topic, helped design the clinical methodology, supervised each step of execution of this study. SM primarily conducted each step of this study ranging from blood sample collection, processing, proteomics experimentation, statistical and proteomic data analysis, results interpretation, and manuscript writing. SSG supervised the proteomics experimentation and its data analysis. SM and PS conducted the proteomics experiments and data analysis. MK, ZR, DB, and RC contributed in conducting the proteomics experimentations in the study. PT and MN contributed in patient sample collection and processing. PK and AK helped in statistical data analysis. DV, PA, AKS, AKP, DM, and KP aided in patient recruitment for this study. All authors contributed to, reviewed, and approved the final draft of the paper.

### Disclosures

The authors report no disclosures relevant to the manuscript.

## Data Availability

All proteomics data produced are available online at ProteomeXchange Consortium via the PRIDE partner repository with the dataset identifier PXD032917.

## Acknowledgement

S. Misra is a DST-INSPIRE Fellow supported by Department of Science and Technology, Government of India.

## Ethics Approval

The study was approved by the local institutional ethics committee of AIIMS, New Delhi (Ref. No. IECPG-395/28.09.2017).

## Glossary

IS: Ischemic Stroke
ICH: Intracerebral hemorrhage
SDE: Significantly Differentially Expressed
CT: Computed Tomography
MRI: Magnetic Resonance Imaging
SWATH-MS: sequential windowed acquisition of all theoretical fragment ion mass spectra
MRM: Multiple Reaction Monitoring
FDR: False Discovery Rate
PCA: Principal Component Analysis
OR: Odds Ratio
CI: Confidence Interval
ROC: Receiver Operating Characteristic
AUC: Area Under the Curve
PPV: Positive Predictive Value
NPV: Negative Predictive Value
VIF: Variance Inflation Factor
IDI: Integrated Discrimination Improvement
GO: Gene Ontology
KEGG: Kyoto Encyclopedia of Genes and Genomes.

