## Supplementary file for "Subtyping strokes using blood-based biomarkers: A proteomics approach"

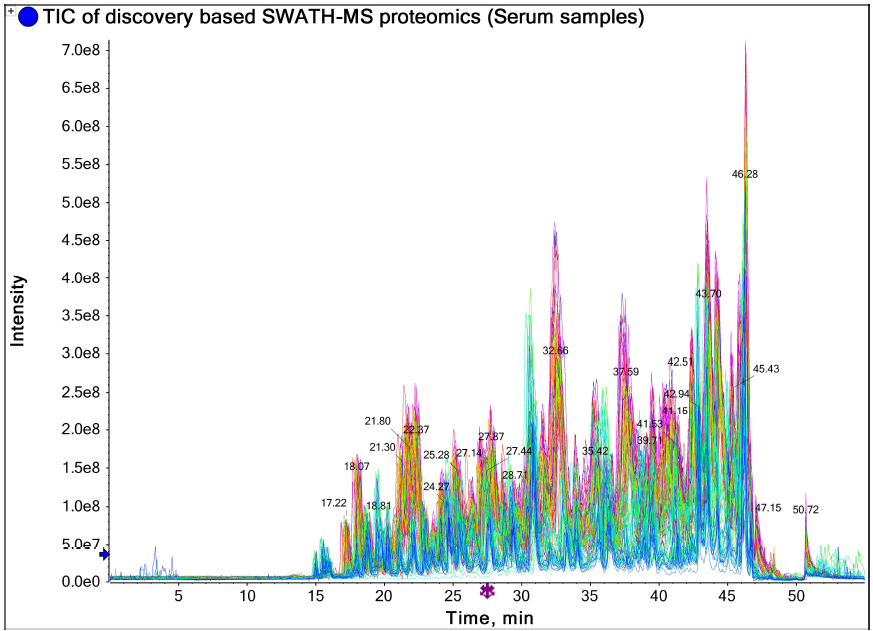

**Supplementary Figure 1:** Total Ion Chromatogram depicting the elution profile of 40 stroke cases and 40 control subjects for the discovery-based SWATH-MS proteomics.

The data on healthy control subjects recruited in the discovery phase of the study is not shown in this paper and is published elsewhere.^1^

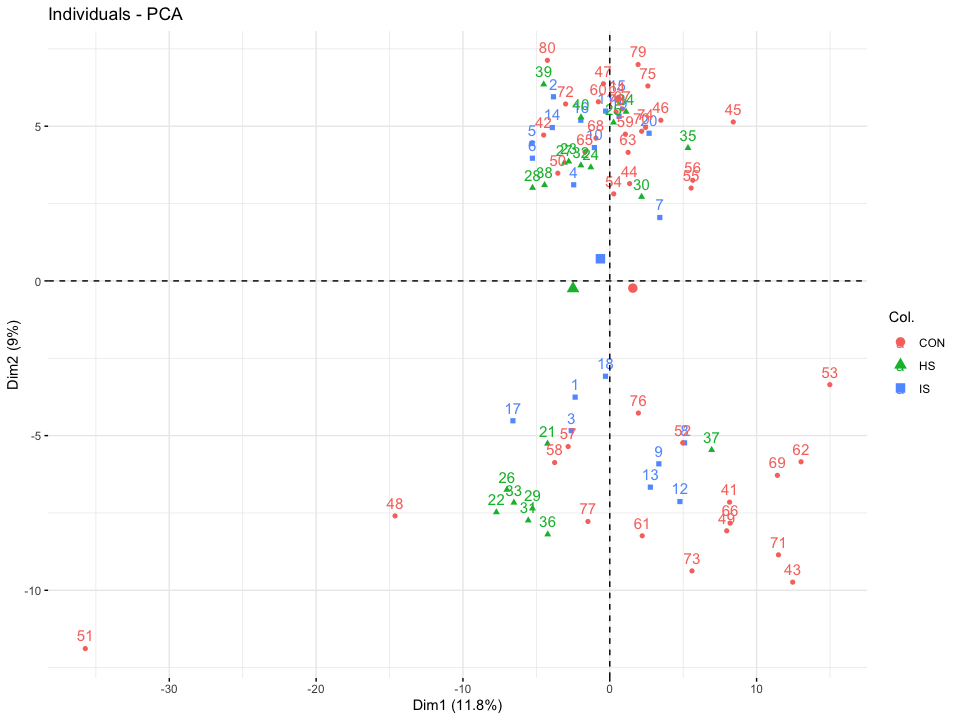

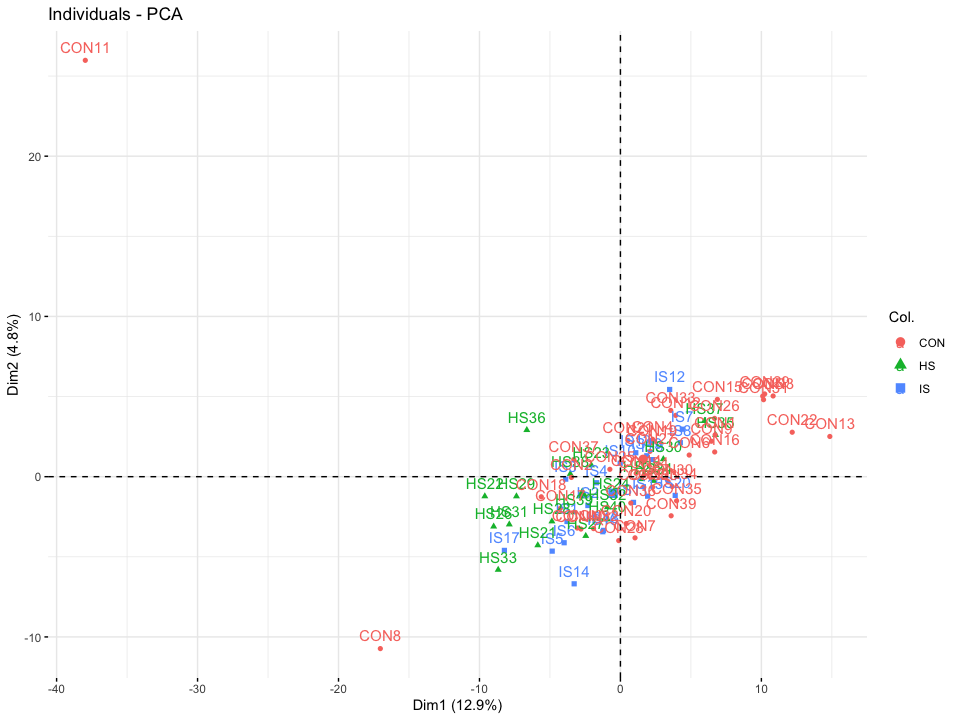
 **Supplementary Figure 2 (a):** Principal Component Analysis plot of the 80 serum samples before batch correction. CON: Healthy Controls; HS: Hemorrhagic Stroke; IS: Ischemic Stroke. **Supplementary Figure 2 (b):** Principal Component Analysis plot of the 80 serum samples after applying the batch correction. CON: Healthy Controls; HS: Hemorrhagic Stroke; IS: Ischemic Stroke.

The data on healthy control subjects recruited in the discovery phase of the study is not shown in this paper and is published elsewhere.^1^

**Supplementary Table 1**: List of shortlisted peptides for MRM

| **S. No** | **Protein Name** | **Peptide Sequence** | **Q1 m/z** | **Q3 m/z** |
| --- | --- | --- | --- | --- |
| 1. | Alpha-2-macroglobulin | AIGYLNTGYQR | 628.3251 | 1071.522  738.3529  851.437  624.31  523.2623  1014.5 |
|  |  | NEDSLVFVQTDK | 697.8435 | 737.3828  836.4512  1151.594  1036.567  949.5353 |
| 2. | Apolipoprotein A-I (APO-A1) | QGLLPVLESFK | 615.8583 | 819.4611  932.5451  623.3399  722.4083  1102.651 |
|  |  | VSFLSALEEYTK | 693.8612 | 940.4622  853.4302  782.3931  1053.546  669.309  540.2664 |
|  |  | DYVSQFEGSALGK | 700.8383 | 1023.511  808.4199  532.3089  661.3515  1122.579  936.4785 |
| 3. | Apolipoprotein L1 (APO-L1) | VNEPSILEMSR | 637.82 | 522.2341  635.3181  748.402  932.487  1061.53 |
| 4. | Polymeric immunoglobulin receptor (PIGR) | TDISMSDFENSR | 467.8699 | 376.1939  505.2365  652.3049  767.3319  854.3639 |
| 5. | Alpha-1-acid glycoprotein 2 (ORM2) | SDVMYTDWK | 572.7526 | 843.3706  712.3301  942.439  1057.466  697.2862 |
|  |  | EHVAHLLFLR | 412.24 | 548.3555  435.271  661.4396  574.2732  366.1772 |
| 6. | Serum amyloid P-component (APCS) | AYSLFSYNTQGR | 703.8386 | 972.4534  825.385  1172.569  1085.537  738.3529 |
| 7. | Apolipoprotein C-I (APO-C1) | TPDVSSALDK | 516.764 | 834.4203  620.325  719.3934  931.4731  533.293 |
|  |  | MREWFSETFQK | 496.9029 | 739.3621  603.2708  652.3301  750.3392  523.2875 |
| 8. | Ig kappa chain V-I region Ni | ASNLETGVPSR | 565.7937 | 745.3839  616.3413  858.468  972.5109  673.3151 |
| 9. | 72 kDa type IV collagenase (MMP2) | VDAAFNWSK | 519.2562 | 823.4097  938.4367  752.3726  681.3355  534.2671 |
| 10. | Multiple inositol polyphosphate phosphatase 1 (MINPP1) | NATALYHVEAFK | 682.3539 | 730.3883  893.4516  1006.536  1077.573  1178.62 |
| 11. | Serpin A11 | SLLHTLALPSPK | 426.259 | 612.3715  541.3344  826.5033  736.4352  849.5193  826.5033 |
| 12. | F-box/WD repeat-containing protein 5 (FBXW5) | TVMVAD**C[CAM]**SR [Carbamidomethyl] | 519.739 | 838.3546  608.2457  707.3141  422.1816  201.1234 |
| 13. | Serotransferrin (TF) | MYLGYEYVTAIR | 739.871 | 1071.547  1184.631  851.4621  1014.525  722.4196 |
|  |  | EGYYGYTGAFR | 642.2882 | 771.3784  934.4417  714.357  1097.505  551.2936 |
|  |  | HSTIFENLANK | 637.3304 | 1136.595  1049.563  829.3839  835.4308  1013.505 |
| 14. | Haptoglobin (HP) | VTSIQDWVQK | 602.322 | 1003.521  803.4046  1104.568  675.3461  916.4887 |
|  |  | TEGDGVYTLNNEK | 720.3361 | 881.4363  1209.575  1037.526  1152.553  980.5047  504.2413  617.3253 |
|  |  | VGYVSGWGR | 490.7511 | 562.2732  881.4264  661.3416  563.2824  824.405 |
| 15. | Beta-2-glycoprotein 1 (APOH) | PDDLPFSTVVPLK | 714.3927 | 987.5873  357.2496  456.318  743.4662  890.5346  656.4341 |
|  |  | VYKPSAGNNSLYR | 490.2563 | 538.2984  451.2663  1078.528  652.3413  1061.501  981.4748 |
|  |  | V**C**[CAM]PFAGILENGAVR  [Cysteine Carbomedomethylated] | 751.8928 | 1243.679  928.5211  999.5582  758.4155  645.3315  516.2889 |
| 16. | Plasma protease C1 inhibitor (SERPING1) | GVTSVSQIFHSPDLAIR | 609.6635 | 908.4948  771.4359  684.4039  1055.563  835.9441 |
|  |  | LLDSLPSDTR | 558.7984 | 575.2784  890.4214  775.3945  557.2678  1003.505  688.3624 |
| 17. | Retinol-binding protein 4 (RBP4) | YWGVASFLQK | 599.8164 | 849.4829  693.393  1035.562  622.3559  792.4614 |
|  |  | DPNGLPPEAQK | 583.296 | 669.3566  572.3039  384.1514  346.2085  839.4621 |
| 18. | Insulin-like growth factor-binding protein 3 (IGFBP3) | FLNVLSPR | 473.2795 | 685.3991  472.2878  571.3562  359.2037  798.4832  375.2027 |
|  |  | EMEDTLNHLK | 615.2952 | 606.2076  969.5  840.4574  725.4304  624.3828  511.2987 |
|  |  | AGASSAGLGPVVR | 571.3198 | 527.33  697.4355  942.5367  470.3085  615.3097  768.4726 |
| 19. | Glial Fibrillary Acidic Protein (GFAP) | HLQEYQDLLNVK | 500.5999 | 701.4192  473.3082  360.2241  812.4512  586.3923 |
|  |  | LEAENNLAAYR | 632.32 | 1021.506  821.4264  950.469  480.2565  707.3855  593.3406 |
|  |  | FADLTDAAAR | 525.7644 | 832.4159  604.3049  717.389  903.453  503.2572  388.2303 |
| 20. | Brain Natriuretic Peptide (BNP) | ISSSSGLGCK | 498.2449 | 882.3986  795.3665  708.3345  621.3025  364.1649 |
|  |  | MVLYTLR | 448.2445 | 665.3802  389.2329  288.1852  764.4487  552.2962 |
| 21. | Matrix metalloprotenase-9 (MMP-9) | AFALWSAVTPLTFTR | 840.959 | 1092.605  835.4672  734.4196  934.5356  403.234  290.1499 |
|  |  | QLSLPETGELDSATLK | 851.4489 | 1260.632  1034.536  933.4888  242.1499  329.1819  519.3137 |
|  |  | AVIDDAFAR | 489.2562 | 464.2616  579.2885  694.3155  807.3995  171.1128 |
| 22. | D-dimer | QGFGNVATNTDGK | 654.8126 | 319.1612  420.2089  635.2995  706.3366  805.405  976.4694 |
|  |  | YLQEIYNSNNQK | 757.3677 | 534.2558  590.2893  704.3322  867.3955  980.4796  1109.522 |
| 23. | Ubiquitin carboxyl-terminal hydrolase isozyme L1 (UCH-L1) | LGVAGQWR | 443.7483 | 546.2783  617.3154  716.3838  773.4053  270.1812  171.1128 |
| 24. | Apolipoprotein A1 (Heavy labelled) (APO-A1 (H)) | QGLLPVLESFK | 619.8654 | 1053.6434  940.5593  827.4753  730.4225  631.3541  518.2700 |

No unique peptide could be identified for Ig heavy chain V-III region VH26 (P01764) and Ig kappa chain V-I region HK101 (P01601) and thus were excluded from the list of selected proteins in the validation phase.

**Abbreviation**: m/z- mass to charge ratio.

**Supplementary Table 2:** Baseline characteristics of acute stroke patients recruited in the discovery phase of the study

| **S. No** | **Characteristics** | **IS patients (N=20)** | **ICH patients (N=20)** | **p-value** |
| --- | --- | --- | --- | --- |
| 1. | Age (years), Mean ± SD & Median (IQR) | 52.85 ± 10.86,  53.5 (45.5-61.5) | 47.60 ± 9.76,  48 (43-55.5) | 0.12 |
| 2. | Male, n (%) | 11 (55) | 14 (70) | 0.33 |
| 3. | Female, n (%) | 9 (45) | 6 (30) |  |
| 4. | Blood sampling time from onset (in hrs.), Mean ± SD & Median (IQR) | 12.11 ± 6.23,  11.5 (7.12-17) | 12.46 ± 6.68,  12.58 (6.25-18.62) | 0.86 |
| 5. | Time taken to reach hospital (in hrs.), Mean ± SD & Median (IQR) | 4.21 ± 2.98,  3.87 (2-5) | 6.41 ± 6.27,  3.75 (2.08-10.12) | 0.16 |
| 6. | Ambulance as a mode of transport, n (%) | 6 (30) | 4 (20) | 0.53 |
| 7. | Any surgical procedure, n (%) | 2 (10) | 5 (25) | 0.21 |
| 8. | Hypertension, n (%) | 8 (40) | 14 (70) | 0.06 |
| 9. | Diabetes, n (%) | 4 (20) | 1 (5) | 0.15 |
| 10. | Dyslipidemia, n (%) | 4 (20) | 0 (0) | **0.03** |
| 11. | Myocardial Infarction, n (%) | 0 | 0 | - |
| 12. | Atrial Fibrillation, n (%) | 0 | 0 | - |
| 13. | Angina Pectoris, n (%) | 1 (5%) | 0 | 0.31 |
| 14. | Migraine, n (%) | 0 | 0 | - |
| 15. | Current Smoking, n (%) | 9 (45) | 10 (50) | 0.75 |
| 17. | Alcohol Intake, n (%) | 2 (10) | 6 (30) | 0.11 |
| 17. | No exercise, n (%) | 18 (90) | 17 (85) | 0.63 |
| 18. | Sedentary lifestyle, n (%) | 7 (35) | 7 (35) | 1.00 |
| 19. | Low Education, n (%) | 13 (65) | 15 (75) | 0.49 |
| 20. | Low socio-economic status, n (%) | 9 (45) | 11 (55) | 0.53 |
| 21. | Obesity, n (%) | 7 (35) | 10 (50) | 0.34 |
| 22. | Family history of stroke, n (%) | 3 (15) | 1 (5) | 0.29 |
| 23. | Family history of hypertension, n (%) | 10 (50) | 6 (30) | 0.20 |
| 24. | Family history of diabetes, n (%) | 7 (35) | 3 (15) | 0.14 |
| 25. | Family history of heart attack, n (%) | 4 (20) | 3 (15) | 0.68 |
| 26. | SBP (mmHg), Mean ± SD & Median (IQR) | 152.7 ± 35.71,  147 (127.5-175) | 178 ± 35.29,  176 (150-214) | **0.03** |
| 27. | DBP (mmHg), Mean ± SD & Median (IQR) | 87.5 ± 17.27,  87 (80-95) | 100.10 ± 17.00,  100 (90-110) | **0.02** |
| 28. | Pulse rate (bpm), Mean ± SD & Median (IQR) | 81.2 ± 13.89,  80 (71-92.5) | 85.38 ± 17.54,  86 (72-97) | 0.42 |

**Abbreviations:** obs: Observations; IS: Ischemic Stroke; ICH: Intracerebral Hemorrhage; SD: Standard Deviation; IQR: Interquartile Range; SBP: Systolic Blood Pressure; DBP- Diastolic Blood Pressure.

**Bold values:** p<0.05.

**Supplementary Table 3**: Results of the confirmed/tentative attributes selected using the Boruta feature selection process between IS and ICH cases within 24 hours

| **Attributes (UniProt ID)** | **Mean Imp** | **Median Imp** | **Min Imp** | **Max Imp** | **Norm Hits** | **Decision** |
| --- | --- | --- | --- | --- | --- | --- |
| P01023 | 4.02 | 4.11 | -0.31 | 6.60 | 0.74 | Confirmed |
| P02787 | 4.10 | 4.14 | -0.24 | 6.69 | 0.75 | Confirmed |
| P04196 | 2.93 | 2.97 | -0.59 | 5.38 | 0.55 | Tentative |
| P02749 | 3.73 | 3.75 | -0.34 | 6.01 | 0.69 | Confirmed |
| P05155 | 5.77 | 5.89 | -0.52 | 8.29 | 0.90 | Confirmed |
| P02753 | 7.42 | 7.60 | 1.46 | 9.87 | 0.97 | Confirmed |
| P01833 | 4.79 | 4.88 | -0.33 | 7.14 | 0.83 | Confirmed |
| P01764 | 5.92 | 6.05 | -0.81 | 8.12 | 0.91 | Confirmed |
| P04431 | 2.49 | 2.57 | -1.00 | 4.86 | 0.45 | Tentative |
| P17936 | 5.85 | 5.97 | 0.56 | 8.19 | 0.91 | Confirmed |
| P02654 | 3.46 | 3.51 | -1.12 | 6.41 | 0.64 | Confirmed |

**Abbreviations**: Imp- Importance measure computed over multiple iterations; Mean Imp- the mean of Imp; Median Imp- the median of Imp; Min Imp- the minimum of Imp; Max Imp- the maximum of Imp; Norm Hits- the number of hits normalized to number of importance source runs.

**Supplementary Table 4:** Functional enrichment analysis of differentially expressed proteins between IS and ICH within 24 hours of symptom onset

| **Category** | **Term name** | **Pathway/process** | **FDR value** | **Proteins involved** | **Background proteins** | **Proteins (gene annotations)** |
| --- | --- | --- | --- | --- | --- | --- |
| **Top 10 processes involved using GO database** | | | | | | |
| GO Cellular Component | GO:0072562 | Blood microparticle | 1.10E-10 | 8 | 115 | APOA1, APCS, SERPING1, APOL1, A2M, HP, TF, ORM2 |
| GO Cellular Component | GO:0005615 | Extracellular space | 4.68E-09 | 16 | 3195 | APOH, MMP2, APOA1, APCS, SERPING1, APOL1, A2M, SERPINA11, HP, PIGR, RBP4, MINPP1, IGFBP3, TF, ORM2, APOC1 |
| GO Cellular Component | GO:0034364 | High-density lipoprotein particle | 5.79E-08 | 5 | 29 | APOH, APOA1, APOL1, HP, APOC1 |
| GO Cellular Component | GO:0034361 | Very-low-density lipoprotein particle | 1.25E-06 | 4 | 20 | APOH, APOA1, APOL1, APOC1 |
| GO Cellular Component | GO:0034774 | Secretory granule lumen | 1.53E-06 | 7 | 324 | APOH, APOA1, SERPING1, A2M, HP, TF, ORM2 |
| GO Cellular Component | GO:0062023 | Collagen-containing extracellular matrix | 4.57E-06 | 7 | 396 | APOH, MMP2, APOA1, APCS, SERPING1, A2M, ORM2 |
| GO Biological Process | GO:0002576 | Platelet degranulation | 1.95E-05 | 6 | 129 | APOH, APOA1, SERPING1, A2M, TF, ORM2 |
| GO Cellular Component | GO:0070062 | Extracellular exosome | 2.59E-05 | 11 | 2099 | APOH, APOA1, APCS, SERPING1, A2M, HP, PIGR, RBP4, MINPP1, TF, ORM2 |
| GO Cellular Component | GO:0030141 | Secretory granule | 2.93E-05 | 8 | 845 | APOH, APOA1, SERPING1, A2M, HP, PIGR, TF, ORM2 |
| GO Cellular Component | GO:0042627 | Chylomicron | 3.91E-05 | 3 | 13 | APOH, APOA1, APOC1 |
| **Pathways involved using KEGG database** | | | | | | |
| KEGG Pathways | hsa04979 | Cholesterol metabolism | 0.005 | 3 | 48 | APOH, APOA1, APOC1 |
| **Top 5 pathways involved using Reactome database** | | | | | | |
| Reactome Pathways | HSA-114608 | Platelet degranulation | 1.77E-05 | 6 | 127 | APOH, APOA1, SERPING1, A2M, TF, ORM2 |
| Reactome Pathways | HSA-381426 | Regulation of Insulin-like Growth Factor (IGF) transport and uptake by Insulin-like Growth Factor Binding Proteins (IGFBPs) | 3.80E-04 | 5 | 124 | MMP2, APOA1, APOL1, IGFBP3, TF |
| Reactome Pathways | HSA-2168880 | Scavenging of heme from plasma | 0.0011 | 3 | 13 | APOA1, APOL1, HP |
| Reactome Pathways | HSA-8963898 | Plasma lipoprotein assembly | 0.0024 | 3 | 19 | APOA1, A2M, APOC1 |
| Reactome Pathways | HSA-8957275 | Post-translational protein phosphorylation | 0.0055 | 4 | 107 | APOA1, APOL1, IGFBP3, TF |

**Abbreviations**: GO- Gene Ontology; KEGG- Kyoto Encyclopedia of Genes and Genomes; FDR- False Discovery Rate.

**Supplementary Table 5**: Baseline characteristics and univariable analysis of ischemic stroke and intracerebral hemorrhage patients recruited within 24 hours in the validation phase of the study

| **S. No** | **Characteristics** | **No. of obs. (IS)** | **IS patients (N=150)** | **No. of obs. (ICH)** | **ICH patients (N=150)** | **OR (95% CI)** | **p-value** | **No. of Obs. (Total Stroke)** | **Total stroke (N=300)** |
| --- | --- | --- | --- | --- | --- | --- | --- | --- | --- |
| 1. | Age (years), Mean ± SD & Median (IQR) | 150 | 54.59 ± 15.54,  55 (44-65) | 150 | 55.30 ± 12.72,  55 (45-65) | 0.99 (0.98-1.01) | 0.67 | 300 | 54.95 ± 14.18,  55 (45-65) |
| 2. | Male, n (%) | 150 | 97 (64.67) | 150 | 98 (65.33) | 1.03 (0.64-1.65) | 0.90 | 300 | 195 (65) |
| 3. | Female, n (%) | 150 | 53 (35.33) | 150 | 52 (34.67) |  |  | 300 | 105 (35) |
| 4. | Blood sampling time from onset (in hrs.), Mean ± SD & Median (IQR) | 150 | 15.29 ± 5.55,  16 (10.5-20.5) | 150 | 14.97 ± 6.55,  17 (10-20) | 1.01 (0.97-1.05) | 0.47 | 300 | 15.13 ± 6.06,  16.37 (10.5-20) |
| 5. | Time taken to reach hospital (in hrs.), Mean ± SD & Median (IQR) | 150 | 5.09 ± 4.61,  3.37 (1.5-7.5) | 150 | 6.17 ± 5.02,  4.71 (2-9.12) | 0.95 (0.91-1.00) | **0.06** | 300 | 5.63 ± 4.84,  4 (2-8.5) |
| 6. | Ambulance as a mode of transport, n (%) | 150 | 27 (18) | 150 | 27 (18) | 1.00 (0.55-1.80) | 1.00 | 300 | 54 (18) |
| 7. | Any surgical procedure, n (%) | 150 | 21 (14) | 150 | 26 (17.33) | 0.78 (0.41-1.45) | 0.43 | 300 | 47 (15.67) |
| **Risk factors for stroke** | | | | | | | | | |
| 8. | Hypertension, n (%) | 150 | 73 (48.67) | 150 | 98 (65.33) | 0.50 (0.32-0.80) | **0.004** | 300 | 171 (57) |
| 9. | Diabetes, n (%) | 150 | 41 (27.33) | 150 | 29 (19.33) | 1.57 (0.91-2.70) | **0.10** | 300 | 70 (23.33) |
| 10. | Dyslipidemia, n (%) | 150 | 16 (10.67) | 150 | 10 (6.67) | 1.67 (0.73-3.81) | 0.22 | 300 | 26 (8.67) |
| 11. | Myocardial Infarction, n (%) | 150 | 6 (4) | 150 | 2 (1.33) | 3.08 (0.61-15.53) | 0.17 | 300 | 8 (2.67) |
| 12. | Atrial Fibrillation, n (%) | 150 | 13 (8.67) | 150 | 3 (2) | 4.65 (1.30-16.67) | **0.02** | 300 | 16 (10.67) |
| 13. | Angina Pectoris, n (%) | 150 | 4 (2.67) | 150 | 1 (0.67) | 4.08 (0.45-36.96) | 0.21 | 300 | 5 (1.67) |
| 14. | Migraine, n (%) | 150 | 1 (0.67) | 150 | 2 (1.33) | 0.50 (0.04-5.53) | 0.57 | 300 | 3 (1) |
| 15. | Current Smoking, n (%) | 150 | 60 (40) | 150 | 41 (27.33) | 1.77 (1.09-2.88) | **0.02** | 300 | 101 (33.67) |
| 16.  16a  16b  16c | Alcohol Intake, n (%) | 150 | 42 (28) | 150 | 35 (23.33) | 1.28 (0.76-2.15) | 0.35 | 300 | 77 (25.67) |
|  | Mild Alcohol, n (%) | 150 | 22 (14.67) | 150 | 13 (8.67) | 1.81 (0.87-3.75) | 0.11 | 300 | 35 (11.67) |
|  | Moderate Alcohol, n (%) | 150 | 5 (3.33) | 150 | 7 (4.67) | 0.70 (0.22-2.27) | 0.56 | 300 | 12 (4) |
|  | Heavy Alcohol, n (%) | 150 | 15 (10) | 150 | 15 (10) | 1.00 (0.47-2.13) | 1.00 | 300 | 30 (10) |
| 17. | No Exercise | 150 | 111 (74) | 150 | 118 (78.67) | 0.77 (0.45-1.32) | 0.34 | 300 | 229 (76.33) |
| 18. | Sedentary lifestyle, n (%) | 111 | 47 (42.34) | 120 | 49 (40.83) | 1.06 (0.63-1.80) | 0.82 | 231 | 96 (41.56) |
| 19. | Low Education, n (%) | 150 | 110 (73.33) | 150 | 115 (76.67) | 0.84 (0.49-1.41) | 0.50 | 300 | 225 (75) |
| 20. | Low socio-economic status, n (%) | 150 | 53 (35.33) | 150 | 52 (34.67) | 1.03 (0.64-1.65) | 0.90 | 300 | 105 (35) |
| 21. | Obesity, n (%) | 150 | 12 (8) | 150 | 18 (12) | 0.64 (0.29-1.37) | 0.25 | 300 | 30 (10) |
| **Family history** | | | | | | | | | |
| 22. | Family history of stroke, n (%) | 150 | 17 (11.33) | 150 | 16 (10.67) | 1.07 (0.52-2.21) | 0.85 | 300 | 33 (11) |
| 23. | Family history of hypertension, n (%) | 150 | 29 (19.33) | 150 | 12 (8) | 1.33 (0.79-2.24) | 0.29 | 300 | 41 (13.67) |
| 24. | Family history of diabetes, n (%) | 150 | 42 (28) | 150 | 34 (22.67) | 2.76 (1.35-5.64) | **0.005** | 300 | 76 (25.33) |
| 25. | Family history of heart attack, n (%) | 150 | 18 (12) | 150 | 9 (6) | 2.14 (0.93-4.92) | **0.07** | 300 | 27 (9) |
| **Vitals at admission** | | | | | | | | | |
| 26. | SBP (mmHg), Mean ± SD & Median (IQR) | 150 | 147.06 ± 33.43,  143 (124-168) | 150 | 175.95 ± 33.29,  171.5 (150-194) | 0.97 (0.96-0.98) | **<0.001** | 300 | 161.50 ± 36.30,  160 (133.5-189) |
| 27. | DBP (mmHg), Mean ± SD & Median (IQR) | 150 | 86.84 ± 18.35,  86 (76-95) | 150 | 100.53 ± 18.82,  100 (88-110) | 0.96 (0.94-0.97) | **<0.001** | 300 | 93.68 ± 19.78,  90 (80-109) |
| 28. | Pulse rate (bpm), Mean ± SD & Median (IQR) | 140 | 84.49 ± 11.72,  84 (79.5-90) | 118 | 85.67 ± 14.03,  84 (78-92) | 0.99 (0.97-1.01) | 0.46 | 258 | 85.03 ± 12.82,  84 (78-90) |
| **Variables at admission and discharge** | | | | | | | | | |
| 29. | BI score at admission, Mean ± SD & Median (IQR) | 150 | 26.07 ± 27.11,  15 (0-45) | 150 | 13.67 ± 23.04,  0 (0-20) | 1.02 (1.01-1.03) | **<0.001** | 300 | 19.87 ± 25.87,  10 (0-35) |
| 30. | NIHSS score at admission, Mean ± SD & Median (IQR) | 150 | 10.94 ± 6.96,  9.5 (6-15) | 150 | 18.25 ± 8.14,  19 (12-26) | 0.89 (0.86-0.92) | **<0.001** | 300 | 14.59 ± 8.40,  13 (8-22) |
| 31. | GCS score at admission, Mean ± SD & Median (IQR) | 150 | 13.57 ± 2.27,  15 (12-15) | 150 | 9.66 ± 4.27,  10 (6-14) | 1.38 (1.27-1.51) | **<0.001** | 300 | 11.62 ± 3.94,  13 (9-15) |
| 32. | mRS score at discharge, Mean ± SD & Median (IQR) | 150 | 3.52 ± 1.55,  4 (3-5) | 150 | 4.39 (1.29),  5 (4-5) | 0.65 (0.54-0.77) | **<0.001** | 300 | 3.96 ± 1.49,  4 (3-5) |
| 33. | Poor outcome at discharge (mRS score 3-6), n (%) | 150 | 113 (75.33) | 150 | 137 (91.33) | 0.29 (0.15-0.57) | **<0.001** | 300 | 250 (83.33) |
| 34. | In-hospital mortality, n (%) | 150 | 7 (4.67) | 150 | 28 (18.67) | 0.21 (0.09-0.50) | **<0.001** | 300 | 35 (11.67) |
| **Blood Investigations** | | | | | | | | | |
| 35. | RBS (mg/dl), Mean ± SD & Median (IQR) | 119 | 151.05 ± 69.25,  127 (110-162) | 111 | 161.58 ± 65.38,  143 (118-187) | 0.99 (0.99-1.00) | 0.24 | 230 | 156.13 ± 67.47,  133 (112-177) |
| 36. | HbA1C (%), Mean ± SD & Median (IQR) | 100 | 6.36 ± 1.76,  5.75 (5.3-6.65) | 49 | 6.11 ± 1.44,  5.7 (5.3-6.4) | 1.10 (0.88-1.37) | 0.39 | 149 | 6.28 ± 1.66,  5.7 (5.3-6.6) |
| 37. | Homocysteine (µM/l), Mean ± SD & Median (IQR) | 88 | 22.45 ± 14.36,  17.76 (13.27-26.92) | 39 | 33.19 ± 22.88,  25.56 (16.42-45.64) | 0.97 (0.95-0.99) | **0.004** | 127 | 25.75 ± 18.03,  20.23 (13.38-31.29) |
| 38. | Total Cholesterol (mg/dl), Mean ± SD & Median (IQR) | 121 | 163.53 ± 48.45,  160 (131-195) | 71 | 163.10 ± 44.71,  154 (133-192) | 1.00 (0.99-1.00) | 0.95 | 192 | 163.37 ± 46.98,  158.5 (132-193) |
| 39. | LDL (mg/dl), Mean ± SD & Median (IQR) | 118 | 107.19 ± 39.65,  108.05 (83-132) | 68 | 99.69 ± 33.18,  98 (74-113.5) | 1.00 (0.99-1.01) | 0.19 | 186 | 104.45 ± 37.50,  102.5 (80-129) |
| 40. | HDL (mg/dl), Mean ± SD & Median (IQR) | 118 | 39.09 ± 11.15,  37 (31-44) | 68 | 40.51 ± 11.89,  38.5 (32.5-46.5) | 0.99 (0.96-1.01) | 0.41 | 186 | 39.61 ± 11.42,  37.5 (32-45) |
| 41. | VLDL (mg/dl), Mean ± SD & Median (IQR) | 118 | 17.58 ± 7.67,  15.5 (12-21) | 68 | 18.07 ± 7.69,  17 (13.5-20.5) | 0.99 (0.95-1.03) | 0.67 | 186 | 17.76 ± 7.66,  16 (12-21) |
| 42. | TG (mg/dl), Mean ± SD & Median (IQR) | 118 | 121.97 ± 59.20,  113 (83-153) | 71 | 146.63 ± 78.21,  122 (97-178) | 0.99 (0.99-0.99) | **0.02** | 189 | 131.24 ± 67.84,  118 (85-156) |
| 43. | Hemoglobin (g/dl), Mean ± SD & Median (IQR) | 136 | 13.23 ± 2.50,  13.25 (11.5-14.8) | 131 | 12.98 ± 2.49,  13.10 (11.2-14.6) | 1.04 (0.94-1.14) | 0.43 | 267 | 13.11 ± 2.49,  13.2 (11.4-14.7) |
| 44. | TLC (10^3^/µL), Mean ± SD & Median (IQR) | 138 | 9161.08 ± 3402.53,  8530 (7200-10500) | 132 | 11198.85 ± 4166.45,  10150 (8790-13000) | 0.99 (0.99-0.99) | **<0.001** | 270 | 10157.32 ± 3923.19,  9310 (7500-12000) |
| 45. | Platelets (10^3^/µL), Mean ± SD & Median (IQR) | 138 | 197.05 ± 92.38,  179 (139-220) | 128 | 186.78 ± 79.87,  183 (120.5-241.5) | 1.00 (0.99-1.00) | 0.33 | 266 | 192.11 ± 86.58,  181.32 (130-228) |
| 46. | Neutrophils (%), Mean ± SD & Median (IQR) | 134 | 68.90 ± 12.19,  70.05 (60.8-78.5) | 86 | 74.89 ± 10.18,  76.35 (70.2-83) | 0.95 (0.93-0.98) | **<0.001** | 220 | 71.24 ± 11.79,  73.9 (63.8-80) |
| 47. | Eosinophils (%), Mean ± SD & Median (IQR) | 134 | 2.15 ± 1.72,  1.6 (1-2.7) | 87 | 1.83 ± 1.99,  1 (0.90-2.2) | 1.11 (0.94-1.31) | 0.20 | 221 | 2.03 ± 1.84,  1.4 (1-2.6) |
| 48. | Basophils (%), Mean ± SD & Median (IQR) | 134 | 1.30 ± 1.77,  0.6 (0.4-1.6) | 89 | 0.92 ± 1.37,  0.5 (0-0.9) | 1.19 (0.97-1.46) | **0.09** | 223 | 1.15 ± 1.63,  0.6 (0.3-1.4) |
| 49. | Lymphocytes (%), Mean ± SD & Median (IQR) | 135 | 22.12 ± 11.04,  20 (12.4-30.3) | 88 | 17.33 ± 9.21,  16.4 (10.85-21.6) | 1.05 (1.02-1.08) | **0.001** | 223 | 20.23 ± 10.60,  18 (12.2-25.8) |
| 50. | Monocytes (%), Mean ± SD & Median (IQR) | 135 | 5.56 ± 2.65,  5.1 (4.1-6.8) | 87 | 4.99 ± 2.65,  4.6 (3-6.8) | 1.09 (0.98-1.21) | 0.12 | 222 | 5.33 ± 2.66,  5 (3.7-6.8) |
| 51. | T3 (ng/dl), Mean ± SD & Median (IQR) | 99 | 87.04 ± 34.06,  88.41 (73.78-105.09) | 34 | 77.02 ± 29.34,  77.39 (59.61-101.71) | 1.01 (0.99-1.02) | 0.13 | 133 | 84.48 ± 33.10,  87.23 (71.41-104.36) |
| 52. | T4 (μg /dl), Mean ± SD & Median (IQR) | 102 | 8.08 ± 8.64,  7.35 (5.9-8.6) | 52 | 8.66 ± 10.03,  7.35 (6-8.8) | 0.99 (0.96-1.03) | 0.71 | 154 | 8.27 ± 9.10,  7.35 (5.9-8.6) |
| 53. | TSH (µIU/ml), Mean ± SD & Median (IQR) | 102 | 5.67 ± 20.63,  2.26 (1.22-3.78) | 57 | 8.64 ± 13.36,  4.1 (0.93-12.2) | 0.99 (0.97-1.01) | 0.35 | 159 | 6.74 ± 18.37,  2.47 (1.08-6.06) |
| 54. | PT (seconds), Mean ± SD & Median (IQR) | 122 | 12.62 ± 1.89,  12.4 (11.6-13) | 75 | 9.64 ± 5.54,  11.7 (2.44-13) | 1.25 (1.13-1.39) | **<0.001** | 197 | 11.49 ± 3.99,  12.2 (11.3-13) |
| 55. | INR, Mean ± SD & Median (IQR) | 120 | 1.20 ± 0.93,  1.10 (1.02-1.14) | 79 | 1.19 ± 0.46,  1.11 (1.03-1.18) | 1.02 (0.70-1.48) | 0.92 | 199 | 1.19 ± 0.78,  1.1 (1.03-1.16) |
| 56. | Vitamin B12 (pg/ml), Mean ± SD & Median (IQR) | 97 | 480.80 ± 484.74,  310 (222-463) | 36 | 578.33 ± 561.58,  327 (237-762) | 0.99 (0.99-1.00) | 0.33 | 133 | 561.58 ± 506.36,  311 (229-486) |
| 57. | Vitamin D (ng/ml), Mean ± SD & Median (IQR) | 43 | 13.33 ± 13.43,  8.9 (5.95-14.91) | 16 | 22.51 ± 24.83,  18.25 (9.54-23.2) | 0.97 (0.94-1.01) | 0.12 | 59 | 15.82 ± 17.53,  10.5 (6.55-20.8) |
| 58. | ESR (mm/hr), Mean ± SD & Median (IQR) | 70 | 23.43 ± 14.53,  22 (12-30) | 23 | 27.95 ± 15.53,  30 (12-38) | 0.98 (0.95-1.01) | 0.21 | 93 | 24.55 ± 14.83,  22 (12-32) |
| 59. | Urea (mg%), Mean ± SD & Median (IQR) | 144 | 30.61 ± 21.68,  26 (20.44-34) | 128 | 38.50 ± 24.85,  29.5 (23-45) | 0.98 (0.97-0.99) | **0.009** | 272 | 34.33 ± 23.52,  27 (21-40) |
| 60. | Creatinine (mg%), Mean ± SD & Median (IQR) | 144 | 0.89 ± 0.76,  0.71 (0.6-1) | 118 | 1.16 ± 0.80,  0.9 (0.7-1.4) | 0.59 (0.39-0.88) | **0.01** | 262 | 1.01 ± 0.79,  0.8 (0.6-1.1) |
| 61. | Potassium (mM/L), Mean ± SD & Median (IQR) | 143 | 4.52 ± 0.71,  4.44 (4.03-4.85) | 127 | 4.08 ± 0.75,  3.9 (3.5-4.5) | 2.42 (1.65-3.56) | **<0.001** | 270 | 4.31 ± 0.76,  4.27 (3.8-4.7) |
| 62. | Sodium (mM/L), Mean ± SD & Median (IQR) | 143 | 140.01 ± 4.70,  140 (137-143.5) | 127 | 138.62 ± 5.84,  139 (135-142) | 1.05 (1.00-1.10) | **0.03** | 270 | 139.35 ± 5.30,  140 (136-142.8) |
| 63. | ALP (I.U.), Mean ± SD & Median (IQR) | 120 | 99.31 ± 37.76,  94 (71.5-111) | 92 | 107.81 ± 40.80,  99.5 (82.5-126) | 0.99 (0.99-1.00) | 0.12 | 212 | 103 ± 39.25,  96.5 (76.5-123) |
| 64. | Calcium (mg%), Mean ± SD & Median (IQR) | 110 | 8.74 ± 0.75,  8.86 (8.4-9.26) | 38 | 8.82 ± 0.66,  8.88 (8.37-9.23) | 0.86 (0.52-1.45) | 0.58 | 148 | 8.76 ± 0.73,  8.87 (8.39-9.25) |
| 65. | SGOT (I.U.), Mean ± SD & Median (IQR) | 117 | 32.30 ± 17.31,  27 (22-39) | 98 | 50 ± 40.85,  37.5 (26-60) | 0.97 (0.96-0.98) | **<0.001** | 215 | 40.37 ± 31.57,  31 (23-45) |
| 66. | SGPT (I.U.), Mean ± SD & Median (IQR) | 117 | 26.46 ± 17.07,  23 (17-30) | 97 | 37.09 ± 32.53,  27 (18-40) | 0.98 (0.97-0.99) | **0.006** | 214 | 31.28 ± 25.77,  24 (17-35) |
| 67. | Total Bilirubin (mg%), Mean ± SD & Median (IQR) | 141 | 0.76 ± 0.62,  0.6 (0.4-0.9) | 116 | 0.90 ± 0.64,  0.7 (0.5-1.05) | 0.70 (0.46-1.06) | **0.09** | 257 | 0.82 ± 0.63,  0.63 (0.49-0.97) |
| 68. | Total Protein (gm%), Mean ± SD & Median (IQR) | 116 | 6.85 ± 0.75,  6.8 (6.5-7.3) | 91 | 6.93 ± 0.97,  7 (6.4-7.6) | 0.89 (0.65-1.24) | 0.50 | 207 | 6.88 ± 0.85,  6.82 (6.4-7.5) |
| 69. | Albumin (gm%), Mean ± SD & Median (IQR) | 116 | 3.79 ± 0.50,  3.9 (3.5-4.2) | 70 | 3.77 ± 0.56,  3.7 (3.4-4.2) | 1.07 (0.61-1.88) | 0.82 | 186 | 3.78 ± 0.52,  3.8 (3.5-4.2) |
| 70. | Globulin (gm%), Mean ± SD & Median (IQR) | 116 | 3.04 ± 0.55,  3 (2.7-3.35) | 69 | 3.18 ± 0.71,  3.2 (2.8-3.6) | 0.69 (0.42-1.14) | 0.15 | 185 | 3.09 ± 0.61,  3.1 (2.7-3.4) |

**Abbreviations:** IS: Ischemic Stroke; ICH: Intracerebral Hemorrhage; OR: Odds Ratio; CI: Confidence Interval; SD: Standard Deviation; IQR: Interquartile range; BI: Barthel Index; NIHSS: National Institutes of Health Stroke Scale; GCS: Glasgow Coma Scale; mRS: modified Rankin Scale; SBP: Systolic Blood Pressure; DBP- Diastolic Blood Pressure; RBS: Random Blood Sugar; LDL: Low Density Lipoprotein; HDL: High Density Lipoprotein; VLDL: Very Low-Density Lipoprotein; TG: Triglyceride; TLC: Total Leucocyte Count; PT: Prothrombin Time; INR: International Normalized Ratio; TSH: Thyroid Stimulating Hormone; ESR: Erythrocyte Sedimentation Rate; SGOT: Serum Glutamic Oxaloacetic Transaminase; SPGT: Serum Glutamic Pyruvic Transaminase; ALP: Alkaline Phosphatase; Obs.: Observations.

**Bold values:** p<0.10.

**Supplementary Table 6**: Baseline characteristics of stroke mimic cases recruited in the validation phase of the study

| **S. No** | **Characteristics** | **Stroke mimics (N=6)** |
| --- | --- | --- |
| 1. | Age (years), Mean ± SD & Median (IQR) | 47.83 ± 18.25  42.5 (34-70) |
| 2. | Male, n (%) | 4 (66.67) |
| 3. | Female, n (%) | 2 (33.33) |
| 4. | Blood sampling time from onset (in hrs.), Mean ± SD & Median (IQR) | 8.75 ± 5.07,  7.25 (6.5-9) |
| 5. | Time taken to reach the hospital (in hrs.), Mean ± SD & Median (IQR) | 4.35 ± 6.50,  1.79 (1-3) |
| **Risk factors for stroke** | | |
| 6. | Hypertension, n (%) | 2 (33.33) |
| 7. | Diabetes, n (%) | 3 (50) |
| 8. | Dyslipidemia, n (%) | 1 (16.67) |
| 9. | Current smoking, n (%) | 1 (16.67) |
| 10. | Alcohol consumption, n (%) | 1 (16.67) |
| 11. | Low education, n (%) | 3 (50) |
| 12. | Low socio-economic status, n (%) | 3 (50) |
| 13. | Family history of stroke, n (%) | 1 (16.67) |
| **Vitals at admission** | | |
| 14. | Systolic Blood Pressure, Mean ± SD & Median (IQR) | 171.6 ± 31.64,  167 (142-198) |
| 15. | Diastolic Blood Pressure, Mean ± SD & Median (IQR) | 96.8 ± 11.69  96 (92-98) |
| **Final diagnosis at discharge** | | |
| 16. | Hypoglycaemia | 2 (33.33) |
| 17. | Vertigo | 2 (33.33) |
| 18. | Syncope | 1 (16.67) |
| 19. | Seizure | 1 (16.67) |
| **Variables at admission and discharge** | | |
| 20. | GCS, Mean ± SD & Median (IQR) | 15 (15-15) |
| 21. | BI at admission, Mean ± SD & Median (IQR) | 73.33 ± 29.61  82.5 (60-95) |
| 22. | NIHSS at admission, Mean ± SD & Median (IQR) | 1.33 ± 1.50  1 (0-2) |
| 23. | mRS at discharge, Mean ± SD & Median (IQR) | 1.67 ± 1.03  1 (1-3) |
| 24. | Poor outcome at discharge (mRS 0-2), n (%) | 2 (33.33%) |
| 25. | Mortality at discharge, n (%) | 0 |

**Abbreviations**: SD: Standard deviation; IQR: Interquartile range; GCS: Glasgow Coma Scale; BI: Barthel Index; NIHSS: National Institutes of Health Stroke Scale; mRS: modified Rankin Scale.

**Supplementary Table 7**: List of peptides and quantifier fragment ion for each protein in the validation phase of the study

| **S. No** | **Protein Name (UniProt ID)** | **Peptide** | **Precursor m/z** | **Product m/z** | **Fragment ion** |
| --- | --- | --- | --- | --- | --- |
| 1. | APO-A1 (P02647) | VSFLSALEEYTK | 693.8612 | 940.462203 | y8 |
| 2. | APO-C1 (P02654) | TPDVSSALDK | 516.764028 | 620.324982 | y6 |
| 3. | APO-L1 (O14791) | VNEPSILEMSR | 637.824094 | 1061.529571 | y9 |
| 4. | APOH (P02749) | VYKPSAGNNSLYR | 490.256285 | 1078.527598 | y10 |
| 5. | MINPP1 (Q9UNW1) | NATALYHVEAFK | 682.353876 | 730.38825 | y6 |
| 6. | ORM2 (P19652) | EHVAHLLFLR | 412.240018 | 435.27143 | y7 |
| 7. | RBP4 (P02753) | YWGVASFLQK | 599.816398 | 849.482879 | y8 |
| 8. | MMP2 (P08253) | VDAAFNWSK | 519.256174 | 823.409714 | y7 |
| 9. | A2M (P01023) | NEDSLVFVQTDK | 697.843538 | 737.382831 | y6 |
| 10. | IGFBP3 (P17936) | EMEDTLNHLK | 615.29517 | 725.43045 | y6 |
| 11. | Haptoglobin (P00738) | TEGDGVYTLNNEK | 720.336078 | 881.436323 | y7 |
| 12. | Serotransferrin (P02787) | MYLGYEYVTAIR | 739.871045 | 1071.546936 | y9 |
| 13. | FBXW5 (Q969U6) | TVMVADCSR | 519.738976 | 422.181629 | y3 |
| 14. | APCS (P02743) | AYSLFSYNTQGR | 703.83859 | 825.384956 | y7 |
| 15. | Serpin A11 (Q86U17) | SLLHTLALPSPK | 426.258967 | 541.334424 | y5 |
| 16. | Ig Kappa Chain V-I region Ni (P01601) | ASNLETGVPSR | 565.793652 | 673.315145 | b7 |
| 17. | SERPING1 (P05155) | LLDSLPSDTR | 558.798403 | 575.278366 | y5 |
| 18. | GFAP (P14136) | HLQEYQDLLNVK | 500.59994 | 586.392273 | y5 |
| 19. | MMP9 (P14780) | AVIDDAFAR | 489.256174 | 807.399543 | y7 |
| 20. | UCH-L1 (P09936) | LGVAGQWR | 443.748319 | 171.112804 | b4 |
| 21. | BNP (P16860) | ISSSSGLGCK | 498.244949 | 882.398557 | y9 |
| 22. | D-dimer | QGFGNVATNTDGK | 654.812573 | 706.336609 | y7 |

**Abbreviations**: APO- Apolipoprotein; APOH: Apolipoprotein H (Beta-2-glycoprotein 1); MINPP1: Multiple inositol polyphosphate phosphatase 1; ORM2: Orosomucoid 2 (Alpha-1-acid glycoprotein 2); RBP4: Retinol Binding Protein 4; MMP- Matrix Metalloproteinase; MMP2: Matrix Metalloproteinase 2 (72 kDa type IV collagenase); A2M: Alpha-2-Macroglobulin; IGFBP3: Insulin-like growth factor-binding protein 3; FBXW5: F-box/WD repeat-containing protein 5; APCS: Serum amyloid P-component; SERPING1: Serpin Family G Member 1 (Plasma protease C1 inhibitor); GFAP: Glial Fibrillary Acidic Protein; UCH-L1: Ubiquitin C-Terminal Hydrolase L1; BNP: Brain Natriuretic Peptide; m/z: mass-to-charge ratio.

**Supplementary Table 8:** Univariable analysis of protein biomarkers assessed in the validation phase of the study to differentiate ischemic stroke and intracerebral hemorrhage within 24 hours of onset

| **S. No** | **Protein biomarker** | **IS patients (N=150)** | **ICH patients (N=150)** | **OR (95% CI)** | **p-value** |
| --- | --- | --- | --- | --- | --- |
| 1. | GFAP, Mean ± SD & Median (IQR) | 14.98 ± 1.30,  15.17 (14.26-15.89) | 16.56 ± 1.10,  16.65 (15.83-17.31) | 0.31 (0.23-0.41) | **<0.001** |
| 2. | MMP9, Mean ± SD & Median (IQR) | 15.50 ± 0.78,  15.63 (15.11-16.01) | 16.04 ± 0.88,  16.01 (15.51-16.61) | 0.44 (0.32-0.60) | **<0.001** |
| 3. | UCH-L1, Mean ± SD & Median (IQR) | 12.77 ± 1.18,  12.94 (12.08-13.59) | 13.36 ± 1.19,  13.57 (12.58-14.16) | 0.66 (0.54-0.81) | **<0.001** |
| 4. | MINPP1, Mean ± SD & Median (IQR) | 19.60 ± 0.64,  19.67 (19.24-20.02) | 19.89 ± 0.70,  19.91 (19.49-20.31) | 0.52 (0.36-0.75) | **<0.001** |
| 5. | APO-A1, Mean ± SD & Median (IQR) | 20.82 ± 0.87,  20.90 (20.19-21.48) | 21.54 ± 0.92,  21.69 (20.88-22.19) | 0.41 (0.30-0.54) | **<0.001** |
| 6. | ORM2, Mean ± SD & Median (IQR) | 20.81 ± 0.79,  20.92 (20.32-21.31) | 21.08 ± 0.71,  21.05 (20.67-21.50) | 0.60 (0.43-0.83) | **0.002** |
| 7. | BNP, Mean ± SD & Median (IQR) (N=179) | 8.92 ± 0.86,  8.91 (8.43-9.46) | 9.29 ± 0.89,  9.36 (8.80-9.73) | 0.59 (0.40-0.87) | **0.008** |
| 8. | APOH, Mean ± SD & Median (IQR) | 19.24 ± 0.75,  19.36 (18.89-19.71) | 19.44 ± 0.79,  19.56 (18.98-19.95) | 0.71 (0.52-0.97) | **0.03** |
| 9. | RBP4, Mean ± SD & Median (IQR) | 16.84 ± 0.81,  16.86 (16.37-17.30) | 17.03 ± 0.84,  17.13 (16.49-17.56) | 0.74 (0.56-0.99) | **0.04** |
| 10. | MMP2, Mean ± SD & Median (IQR) | 18.79 ± 0.62,  18.79 (18.42-19.20) | 18.90 ± 0.71,  18.91 (18.55-19.27) | 0.78 (0.55-1.10) | **0.07** |
| 11. | A2M, Mean ± SD & Median (IQR) | 18.20 ± 0.76,  18.29 (17.79-18.70) | 18.34 ± 0.88,  18.39 (17.91-18.97) | 0.81 (0.61-1.08) | **0.08** |
| 12. | APO-C1, Mean ± SD & Median (IQR) | 16.87 ± 1.20,  17.04 (16.40-17.55) | 16.66 ± 1.36,  16.81 (15.92-17.64) | 1.14 (0.95-1.36) | **0.09** |
| 13. | Haptoglobin, Mean ± SD & Median (IQR) | 18.26 ± 1.23,  18.54 (17.67-19.02) | 17.98 ± 1.58,  18.19 (17.22-18.98) | 1.15 (0.98-1.36) | **0.09** |
| 14. | Serotransferrin, Mean ± SD & Median (IQR) | 20.13 ± 0.76,  20.10 (19.74-20.62) | 20.28 ± 0.83,  20.33 (19.76-20.80) | 0.79 (0.59-1.05) | **0.09** |
| 15. | IGFBP3, Mean ± SD & Median (IQR) | 16.43 ± 0.73,  16.43 (16.02-16.91) | 16.57 ± 0.78,  16.51 (16.14-17.10) | 0.78 (0.57-1.05) | **0.10** |
| 16. | FBXW5, Mean ± SD & Median (IQR) (N=175) | 7.77 ± 1.12,  7.70 (7.00-8.66) | 8.03 ± 1.07,  8.08 (7.25-8.77) | 0.81 (0.61-1.06) | 0.13 |
| 17. | APCS, Mean ± SD & Median (IQR) (N=229) | 14.69 ± 1.68,  15.14 (13.42-15.93) | 14.98 ± 1.78,  15.44 (14.01-16.22) | 0.91 (0.78-1.06) | 0.21 |
| 18. | APO-L1, Mean ± SD & Median (IQR) | 19.45 ± 0.79,  19.52 (18.96-20.02) | 19.53 ± 0.89,  19.54 (18.97-20.24) | 0.89 (0.68-1.16) | 0.39 |
| 19. | Ig Kappa Chain, Mean ± SD & Median (IQR) | 16.14 ± 1.10,  16.29 (15.54-16.85) | 16.23 ± 1.09,  16.28 (15.64-17.06) | 0.93 (0.75-1.14) | 0.48 |
| 20. | SERPING 1, Mean ± SD & Median (IQR) | 18.26 ± 1.16,  18.53 (17.90-18.99) | 18.35 ± 1.11,  18.59 (18.02-18.97) | 0.93 (0.76-1.14) | 0.48 |
| 21. | D-dimer, Mean ± SD & Median (IQR) (N=73) | 11.92 ± 2.84,  11.93 (10.19-13.34) | 12.35 ± 2.88,  11.80 (10.38-13.65) | 0.95 (0.80-1.12) | 0.52 |
| 22. | Serpin A11, Mean ± SD & Median (IQR) | 13.35 ± 1.16,  13.43 (12.77-14.05) | 13.42 ± 1.17,  13.49 (12.80-14.20) | 0.95 (0.78-1.16) | 0.61 |

**Abbreviations**: OR: Odds Ratio; CI: Confidence Interval; APO- Apolipoprotein; APOH: Apolipoprotein H (Beta-2-glycoprotein 1); MINPP1: Multiple inositol polyphosphate phosphatase 1; ORM2: Orosomucoid 2 (Alpha-1-acid glycoprotein 2); RBP4: Retinol Binding Protein 4; MMP- Matrix Metalloproteinase; MMP2: Matrix Metalloproteinase 2 (72 kDa type IV collagenase); A2M: Alpha-2-Macroglobulin; IGFBP3: Insulin-like growth factor-binding protein 3; FBXW5: F-box/WD repeat-containing protein 5; APCS: Serum amyloid P-component; SERPING1: Serpin Family G Member 1 (Plasma protease C1 inhibitor); GFAP: Glial Fibrillary Acidic Protein; UCH-L1: Ubiquitin C-Terminal Hydrolase L1; BNP: Brain Natriuretic Peptide.

**Bold values:** p<0.10.

**Supplementary Table 9**: Baseline characteristics and univariable analysis of ischemic stroke and intracerebral hemorrhage patients recruited within 6 hours in the validation phase of the study

| **S. No** | **Characteristics** | **IS patients (N=12)** | **ICH patients (N=38)** | **OR (95% CI)** | **p-value** |
| --- | --- | --- | --- | --- | --- |
| 1. | Age (years), Mean ± SD & Median (IQR) | 58.92 ± 13.38  62 (48.5-68.5) | 55.65 ± 12.86  58.5 (49-66) | 1.02 (0.96-1.08) | 0.67 |
| 2. | Male, n (%) | 7 (58.33) | 22 (84.62) | 3.93 (0.82-18.81) | **0.09** |
| 3. | Female, n (%) | 5 (41.67) | 4 (15.38) |  |  |
| 4. | Blood sampling time from onset (in hrs.), Mean ± SD & Median (IQR) | 5.00 ± 0.92  4.87 (4.5-6) | 3.93 ± 1.44  4 (3.25-5.3) | 2.13 (1.05-4.31) | **0.03** |
| 5. | Time taken to reach hospital (in hrs.), Mean ± SD & Median (IQR) | 2.66 ± 1.22  2.5 (2-3) | 3.53 ± 3.47  2 (1.5-4.5) | 0.88 (0.66-1.18) | 0.40 |
| 6. | Ambulance as a mode of transport, n (%) | 2 (16.67) | 3 (11.54) | 1.53 (0.22-10.64) | 0.66 |
| 7. | Any surgical procedure, n (%) | 1 (8.33) | 1 (3.85) | 2.27 (0.13-39.73) | 0.57 |
| **Risk factors for stroke** | | | | | |
| 8. | Hypertension, n (%) | 4 (33.33) | 14 (53.85) | 0.43 (0.10-1.78) | 0.24 |
| 9. | Diabetes, n (%) | 3 (25) | 6 (23.08) | 1.11 (0.22-5.47) | 0.90 |
| 10. | Dyslipidemia, n (%) | 2 (16.67) | 1 (3.85) | 5.00 (0.41-61.52) | 0.21 |
| 11. | Current Smoking, n (%) | 4 (33.33) | 5 (19.23) | 2.10 (0.45-9.86) | 0.35 |
| 12. | Alcohol Intake, n (%) | 3 (25) | 9 (34.62) | 0.63 (0.13-2.93) | 0.55 |
| 13. | No Exercise | 7 (58.33) | 21 (80.77) | 0.33 (0.07-1.50) | 0.15 |
| 14. | Low Education, n (%) | 10 (83.33) | 20 (76.92) | 1.5 (0.25-8.82) | 0.65 |
| 15. | Low socio-economic status, n (%) | 4 (33.33) | 8 (30.77) | 1.12 (0.26-4.85) | 0.87 |
| 16. | Family history of hypertension, n (%) | 2 (16.67) | 8 (30.77) | 0.45 (0.08-2.54) | 0.37 |
| 17. | Family history of diabetes, n (%) | 1 (8.33) | 4 (15.38) | 0.50 (0.05-5.02) | 0.56 |
| 18. | Family history of heart attack, n (%) | 1 (8.33) | 2 (7.69) | 1.09 (0.09-13.35) | 0.95 |
| **Vitals at admission** | | | | | |
| 19. | Systolic blood pressure, (Mean ± SD) | 141 ± 29.58  135.5 (120-159) | 173 ± 26.64  170 (158-190) | 0.96 (0.93-0.99) | **0.008** |
| 20. | Diastolic blood pressure, (Mean ± SD) | 76.58 ± 11.88  80 (64.5-86) | 99.15 ± 16.42  100 (90-110) | 0.89 (0.83-0.96) | **0.004** |
| **Variables at admission and discharge** | | | | | |
| 21. | BI score at admission, Mean ± SD & Median (IQR) | 35.83 ± 34.37  25 (15-57.5) | 17.88 ± 23.71  0 (0-45) | 1.02 (0.99-1.05) | **0.08** |
| 22. | NIHSS score at admission, Mean ± SD & Median (IQR) | 8.33 ± 7.73  6.5 (3.5-11.5) | 15.27 ± 6.96  13 (11-22) | 0.86 (0.76-0.98) | **0.02** |
| 23. | GCS score at admission, Mean ± SD & Median (IQR) | 13.92 ± 2.06  15 (13.5-15) | 11.11 ± 4.10  13 (8-15) | 1.37 (0.99-1.88) | **0.06** |
| 24. | mRS score at discharge, Mean ± SD & Median (IQR) | 2.75 ± 1.81  4 (1-4) | 4.35 ± 1.26  4.5 (4-5) | 0.50 (0.29-0.85) | **0.01** |
| 25. | Poor outcome at discharge (mRS score 3-6), n (%) | 7 (58.33) | 23 (88.46) | 0.18 (0.03-0.96) | **0.04** |
| 26. | In-hospital mortality, n (%) | 0 | 5 (19.23) | **-** | |

**Abbreviations:** IS: Ischemic Stroke; ICH: Intracerebral Hemorrhage; OR: Odds Ratio; CI: Confidence Interval; SD: Standard Deviation; IQR: Interquartile Range; BI: Barthel Index; NIHSS: National Institutes of Health Stroke Scale; GCS: Glasgow Coma Scale; mRS: modified Rankin Scale.

**Bold values:** p<0.10.

**Supplementary Table 10**: The diagnostic potential of protein biomarkers assessed in the validation phase of the study to differentiate ischemic stroke and intracerebral hemorrhage within 6 hours of onset

| **S. No** | **Protein Biomarker (UniProt ID)** | **OR (95% CI)** | **p-value** | **Cutoff** | **Sensitivity (95% CI)** | **Specificity (95% CI)** | **PPV (95% CI)**  **(For IS)** | **NPV (95% CI)**  **(For ICH)** |
| --- | --- | --- | --- | --- | --- | --- | --- | --- |
| 1 | GFAP (P14136) | 0.02 (0.002-0.17) | <0.001 | <15.55 | 92% (62-99%) | 85% (65-96%) | 73% (45-92%) | 95% (78-99%) |
| 2 | Alpha-2-Macroglobulin (P01023) | 0.15 (0.03-0.68) | 0.01 | <17.86 | 67% (35-90) | 77% (56-91%) | 57% (29-82%) | 83% (63-95%) |
| 3 | IGFBP3 (P17936) | 0.10 (0.20-0.59) | 0.01 | <16.38 | 83% (52-98%) | 65% (44-83%) | 53% (29-76%) | 89% (67-99%) |
| 4 | Serpin A11 (Q86U17) | 0.18 (0.04-0.86) | 0.03 | <12.72 | 50% (21-79%) | 85% (65-96%) | 60% (26-88%) | 79% (59-92%) |
| 5 | APO-A1 (P02647) | 0.09 (0.01-0.81) | 0.03 | <21.50 | 92% (61-99%) | 50% (30-70%) | 46% (26-67%) | 93% (66-99%) |
| 6 | BNP (P16860) (N=25) | 0.13 (0.02-0.86) | 0.03 | <8.79 | 62% (25-92%) | 82% (57-96%) | 62% (25-92%) | 82% (57-96%) |
| 7 | MMP9 (P14780) | 0.11 (0.01-0.94) | 0.04 | <16.05 | 92% (61-99%) | 46% (27-67%) | 44% (24-65%) | 92% (64-99%) |
| 8 | RBP4 (P02753) | 0.17 (0.03-0.94) | 0.04 | <17.11 | 83% (52-98%) | 54% (33-73%) | 45% (24-68%) | 88% (62-98%) |
| 9 | FBXW5 (Q969U6)  (N=24) | 0.15 (0.02-1.00) | 0.05 | <7.54 | 75% (35-97%) | 69% (41-89%) | 55% (23-83%) | 85% (55-98%) |

The cut-off values represent the Log_2_ normalized protein concentrations.

**Abbreviations**: OR: Odds Ratio; CI: Confidence Interval; PPV: Positive Predictive Value; NPV: Negative Predictive Value; APO- Apolipoprotein; APOH: Apolipoprotein H (Beta-2-glycoprotein 1); MINPP1: Multiple inositol polyphosphate phosphatase 1; ORM2: Orosomucoid 2 (Alpha-1-acid glycoprotein 2); RBP4: Retinol Binding Protein 4; MMP- Matrix Metalloproteinase; MMP2: Matrix Metalloproteinase 2 (72 kDa type IV collagenase); A2M: Alpha-2-Macroglobulin; IGFBP3: Insulin-like growth factor-binding protein 3; FBXW5: F-box/WD repeat-containing protein 5; APCS: Serum amyloid P-component; SERPING1: Serpin Family G Member 1 (Plasma protease C1 inhibitor); GFAP: Glial Fibrillary Acidic Protein; UCH-L1: Ubiquitin C-Terminal Hydrolase L1; BNP: Brain Natriuretic Peptide.

**Supplementary Table 11**: The diagnostic potential of protein biomarkers assessed in the validation phase of the study to differentiate ischemic stroke and stroke mimics within 24 hours of onset

| **S. No** | **Protein Biomarker (UniProt ID)** | **OR (95% CI)** | **p-value** | **Cutoff** | **Sensitivity** | **Specificity** | **PPV**  **(For IS)** | **NPV**  **(For Mimics)** |
| --- | --- | --- | --- | --- | --- | --- | --- | --- |
| 1 | APO-L1 (O14791) | 18.43 (2.07-163.47) | 0.009 | >18.86 | 79% (71-85%) | 83% (36-99%) | 99% (95-99%) | 13% (4-29%) |
| 2 | BNP (N=96) (P16860) | 0.08 (0.009-0.77) | 0.03 | <9.35 | 70% (59-79%) | 83% (36-99%) | 98% (92-99%) | 16% (5-33%) |
| 3 | FBXW5 (Q969U6)  (N=95) | 0.11 (0.01-1.00) | 0.05 | <8.25 | 64% (53-74%) | 83% (36-99%) | 98% (91-99%) | 13% (4-29%) |

The cut-off values represent the Log2 normalized protein concentrations.

**Abbreviations**: OR: Odds Ratio; CI: Confidence Interval; PPV: Positive Predictive Value; NPV: Negative Predictive Value; APO- Apolipoprotein; FBXW5: F-box/WD repeat-containing protein 5; BNP: Brain Natriuretic Peptide.
